# Association of complex traits with common genetic variation across genomic regions containing pathogenic copy number variations

**DOI:** 10.1101/2024.09.18.24313729

**Authors:** Yelyzaveta Snihirova, Esmee M. Breddels, Oleksandr Frei, Ida E. Sønderby, Ole A Andreassen, Therese van Amelsvoort, David E.J. Linden, Dennis van der Meer

**Author notes:** **Corresponding author(s):** Dr. Dennis van der Meer., Yelyzaveta Snihirova.

## Abstract

**Background:** Copy Number Variations (CNVs) are structural variation in the genome, which may impact complex human traits and diseases. The investigation of rare CNVs is impeded by low sample size. To understand the mechanisms through which CNVs influence human health, common variation in the genomic region of the CNV from large samples could be used as a proxy.

**Methods:** Utilising genome-wide association study (GWAS) summary statistics of 20 traits, we assessed the cumulative effect of common genetic variants in eight genomic regions containing pathogenic CNVs, using MAGMA gene-based analysis. We used GSA-MiXeR to estimate the fold enrichment of these CNV regions for the specific phenotypes.

**Results:** The distal and proximal regions of the 16p11.2 CNV exhibited the highest number of significant associations and were enriched for the highest number of traits: 12 of 27 significant MAGMA associations (44%) were enriched. These CNV regions also had the highest number of phenotype-associated genes related to ion transport, signalling, transcriptional regulation, development, and protein metabolism. We compared the significance of all the genomic regions and the genes in these regions and found two opposing patterns: 1) cumulative value of separate genes, resulting in the higher significance of the whole region than of the particular genes; 2) higher significance of the specific genes that drive the association of the whole region.

**Conclusions:** Charting the features of genomic regions encompassing CNVs might aid in clarifying CNVs’ role in human disease, especially pinpointing candidate genes within these regions that are associated with complex traits.

## Introduction

Copy number variations (CNVs) are DNA segments where the count of specific sequences varies among individuals. CNVs arise due to deletions or duplications of these segments (1). They span 12-16% of the human genome (2) and typically occur in certain susceptible areas of the genome, which are characterized by instability, segmental duplications, low-copy repeats, and propensity to genomic rearrangements (3,4).

Rare, pathogenic CNVs are associated with the development of complex human diseases and traits (1), particularly in the area of neurodevelopment (5). The main mechanisms by which they exert a pathogenic effect are altering gene expression and regulation (6,7) and causing genomic instability (1,8) Polygenic complex traits associated with CNVs include mental disorders (9), cognition (10), anthropomorphic measures (11), blood biomarkers (12), congenital heart disease (13), and cancer (14).

The study of the effects of CNVs on human health faces limitations. Many CNVs remain undetected or unreported, often due to a lack of testing or absence of clinical symptoms in individuals (15). The different CNV detection methods vary in specificity and sensitivity, posing challenges for standardization (16,17). While common CNVs are relatively frequent and may be present in a substantial proportion of the population, rare CNVs are often more relevant to disease-related studies (18). Consequently, sample sizes in CNV research in human diseases are often small, making studies underpowered. Unlike CNVs, information on common genetic variants in the human genome and their effect on human health is widely available through summary statistics from large genome-wide association studies (GWAS) (19).

This study seeks to elucidate the relationship between complex traits and common genetic variation within genomic regions containing rare pathogenic CNV. The regions were confined to those including eight known and common pathogenic CNVs. We analyzed 20 complex human traits in six categories: mental disorders, substance use, cognition, somatic disorders, blood biomarkers, and anthropometrics. This allowed us to assess associations of the common genetic variation in CNV regions with individual phenotypes and examine association patterns within and across categories. We postulate that aggregating the effects of common genetic variation within the genomic region of a CNV may be used as a proxy to study how that CNV affects human health. The large benefit of this approach is the large increase in sample size, as the current generation of GWAS of complex traits is often based on tens or hundreds of thousands of individuals. Further, this approach allows us to study associations between specific sub-regions and genes in the genomic region with traits of interest, unveiling new insights into the influence of CNVs on these traits.

## Methods and Materials

All analyses were conducted in R version 4.2.3, with quality control procedures for GWAS summary statistics implemented in Python versions 3.7 and 2.7.

### Genomic regions of interest

This study focused on specific genomic regions containing pathogenic CNVs, namely 1q21.1 distal (1q21.1d), 3q29, 7q11.23, 15q11.2 BP1-BP2, 15q13.3, 16p11.2 distal (16p11.2d), 16p11.2 proximal (16p11.2p), and 22q11.2. These regions were chosen for their known associations with schizophrenia or severe neurodevelopmental phenotypes, as reported by Kirov et al. (20), and are among the most common pathogenic CNV loci (4). Genomic coordinates were extracted from Ehrhart et al. (21) and Kendall et al. (10), originally obtained via BioMart query (GRCh37/hg19), and are listed in Table 1. Due to the limited data on the prevalence of CNVs in the general population (22), we included the number of carriers from the UK Biobank population cohort according to Owen et al. (23), as presented in Table 1. This provides an estimate of the sample size (ranging from 1 to 3067 out of approximately 400 000) if carrier status was studied in such a cohort, demonstrating the value of our current approach.

**Table 1.** Summary of genomic regions of interest used in the study sorted in decreasing order according to their size. Start and end position and length are given in base pairs. Location is based on human genome GRCh37/hg19.

| Genomic region | Chromosome | Start position | End position | Length | Number of carriers in UK Biobank (23) |
| --- | --- | --- | --- | --- | --- |
| 22q11.2 | 22 | 18912231 | 21465672 | 2 553 441 | 22q11.2 del – 10<br>22q11.2 dup – 266 |
| 15q13.3 | 15 | 30500000 | 32500000 | 2 000 000 | 15q13.3 del – 37<br>15q13.3 dup – 224 |
| 7q11.23 | 7 | 72744454 | 74142513 | 1 398 059 | 7q11.23 del (Williams-Beuren syndrome (WBS) del) – 1<br>7q11.23 dup (WBS dup) – 13 |
| 3q29 | 3 | 195788299 | 197033296 | 1 244 997 | 3q29 del – 9<br>3q29 dup – 5 |
| 1q21.1d | 1 | 146527987 | 147394444 | 866 457 | 1q21.1 del – 106<br>1q21.1dup – 168 |
| 16p11.2p | 16 | 29592751 | 30190593 | 597 842 | 16p11.2p del – 103<br>16p11.2p dup – 131 |
| 15q11.2 BP1-BP2 | 15 | 22805313 | 23094530 | 289 217 | 15q11.2 BP1-BP2 del – 1570<br>15q11.2 BP1-BP2 dup – 1917 |
| 16p11.2d | 16 | 28823196 | 29046783 | 223 587 | 16p11.2d del – 54<br>16p11.2d dup – 127 |

### Complex traits included

We investigated the genetic association between the listed genomic regions of interest and various complex traits using summary statistics obtained from previously published GWAS:

a. Mental disorders (attention deficit hyperactivity disorder (ADHD) (24), Alzheimer’s disease (25), autism spectrum disorder (ASD) (26), bipolar disorder (27), major depressive disorder (MDD) (28), post-traumatic stress disorder (PTSD) (29), schizophrenia (30));
b. Substance use (Alcohol Use Disorders Identification Test (AUDIT score) (31), cannabis use (32));
c. Cognition (educational attainment (33), intelligence (34));
d. Somatic disorders (coronary artery disease (35), chronic kidney disease (36), type II diabetes (T2D) (37), lung cancer (38));
e. Blood biomarkers (white blood cell count (WBC) (39), high-density lipoprotein in blood (HDL) (40), calcium in blood (41));
f. Anthropometrics (height (11), body mass index (BMI) (42)).

### Region- and gene-based analysis of association between common variants and traits using MAGMA

To assess the collective impact of multiple SNPs within and across specific genomic regions, we employed gene-based analyses through Multi-marker Analysis of GenoMic Annotation (MAGMA v1.09) (43). MAGMA utilizes GWAS summary statistics to aggregate effects of common variants within a genomic region and calculates p-values using an F-test for region-phenotype associations. We derived aggregated effects and corresponding p-values for each combination of genomic region and phenotype, considering linkage disequilibrium using Phase 3 data from the 1000 Genomes European population. Bonferroni correction was applied to MAGMA results to correct for multiple comparisons, setting the threshold at alpha=.05/20*8 (8 chromosomal regions × 20 traits) = 3.1*10^−4^.

Furthermore, we utilized MAGMA to investigate individual genes within our regions of interest, identifying genes responsible for driving significant associations between genomic regions and specific phenotypes. This approach allowed us to distinguish between trait-specific genes and those exhibiting significance across multiple traits, offering insights into shared genetic pathways underlying diverse phenotypic traits. We employed NCBI 37.3 gene definitions within relevant genomic boundaries, aggregating effects of all common variants within each gene to calculate p-values for gene-trait associations. To compare significance between entire region and individual genes, we examined if the region’s significance surpassed the 1.96× standard deviation (SD) cut-off of the gene-level significance distribution.

### Measures of genetic architecture using MiXeR

The polygenicity of the trait influences the likelihood of detecting significant differences between the genomic region of interest and random regions of equal size. To estimate the polygenicity of each trait, representing the number of causal variants explaining 90% of SNP heritability, we utilized MiXeR v1.3 (44,45). This provided insight into the identified association patterns: genomic regions of interest are less likely to differ significantly from random regions of the same size in terms of trait association under higher polygenicity.

As part of MiXeR quality control, we excluded variants within the major histocompatibility complex (MHC) region (GRCh37: 6:26MB – 34MB), filtered SNPs with odds ratios exceeding a 1×10^37^ threshold (to remove SNPs with excessively large odds, likely artefacts of outliers), and retained only HapMap3 SNPs in GWAS summary statistics. Additionally, we extracted the traits’ discoverability (effect size variance) and SNP-based heritability (the degree of variation in a phenotypic trait explained by SNPs).

### Partitioned heritability and enrichment via GSA-MiXeR

To complement MAGMA results, we utilised GSA-MiXeR (46), a competitive gene-set enrichment analysis method. GSA-MiXeR assesses partitioned local heritability, indicating how much of the SNP-based heritability is explained by a specific genomic region, along with fold enrichment of genes and gene sets, reflecting how strongly a region is associated with a phenotype trait compared to chance. We employed GSA-MiXeR to evaluate the contribution of the genomic region of interest to SNP-heritability of polygenic traits, considering factors such as polygenicity, gene size, minor allele frequency (MAF), and LD-dependent genetic architecture (46). Furthermore, we aimed to assess the collective contribution of all genomic regions to the phenotype trait, evaluating the enrichment of all CNV-containing regions.

We employed Frei et al.’s framework (46) for analysis, treating genomic regions of interest as ‘genes’ and comparing them to the whole genome as an infinitesimal baseline model. Reference data were sourced from Phase 3 data of the 1000 Genomes European population and the GRCh37/hg19 genomic build. We extracted the mean estimated heritability (indicating the extent of phenotypic trait variation attributed to the region of interest) and mean enrichment (the ratio of heritability estimates compared to the baseline infinitesimal model), alongside their standard deviations from 20 runs of the GSA-MiXeR model. Each run was constrained to a random subset of genetic variants during the fitting procedure. We classified a region as “enriched” for the phenotype trait if its mean of enrichment-1.96×SD exceeded 1.

As a robustness analysis, we compared the partitioned heritability estimate from GSA-MiXeR with another tool, LDAK-GBAT (47). LDAK-GBAT is a gene-based association testing tool that estimates local heritability via permutations using the UK Biobank reference panel.

We assessed correlations and differences between MAGMA, GSA-MiXeR, and LDAK-GBAT results, as well as their correlations with region sizes and trait polygenicity, using Spearman’s rank correlation coefficient and t-statistic.

## Results

### Aggregation of common variants across the genomic regions encompassing CNVs

We utilized MAGMA to evaluate the association between common variants in eight distinct regions of interest and 20 traits (Table S1 and Figure 1).

**Figure 1.**
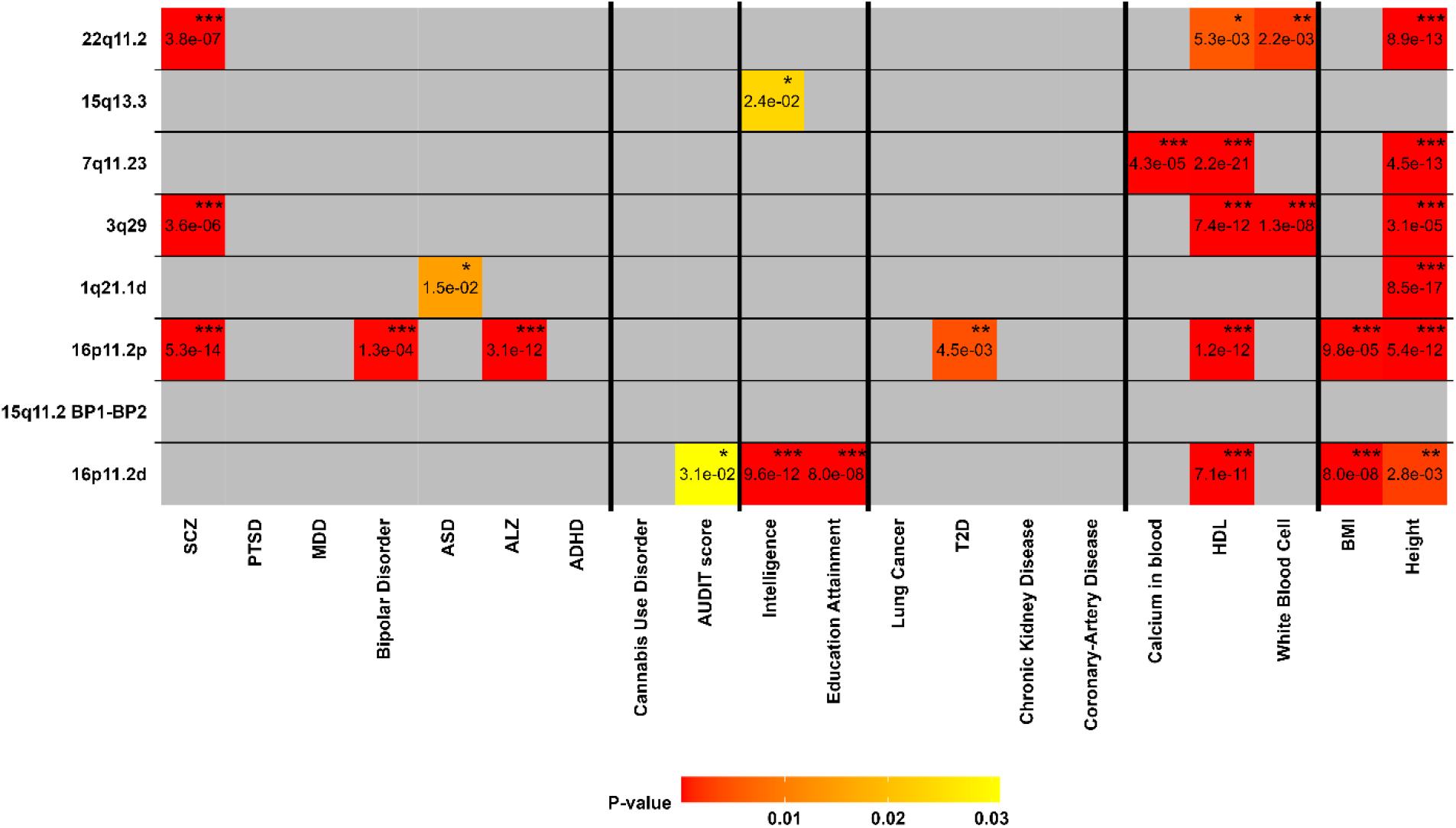
MAGMA-based gene analysis for complex traits and disorders. Colour denotes significant trait-region of interest associations with significance levels expressed by asterisks (*** for p-value ≤ 0.0005, ** for p-value ≤ 0.005, * for p-value ≤ 0.05; Bonferroni-corrected). Regions of interest are sorted in size-decreasing order on the x-axis, and traits are categorised on the y-axis. ADHD = Attention Deficit Hyperactivity Disorder, AUDIT score = Alcohol Use Disorders Identification Test (AUDIT) score, ALZ = Alzheimer’s disease, ASD = Autism Spectrum Disorder, BMI = Body Mass Index, HDL = High-Density Lipoprotein, MDD = Major Depressive Disorder, PTSD = Post-Traumatic Stress Disorder, SCZ = Schizophrenia, T2D = Type II Diabetes.

The 16p11.2d and 16p11.2p regions demonstrated the highest number of significant associations, six and seven, respectively. Both regions were associated with HDL, BMI, and height. Moreover, 16p11.2p displayed associations with T2D and multiple mental disorders (schizophrenia, bipolar disorder, and Alzheimer’s disease), while 16p11.2d was linked to the AUDIT score and cognition traits. In contrast, 15q11.2 BP1-BP2 did not exhibit significant associations with any traits. Both the 3q29 and 22q11.2 regions were associated with similar phenotypes, including schizophrenia, HDL in blood, WBC, and height. Regarding association patterns between trait categories, 16p11.2d showed significant associations with cognition (intelligence and educational attainment) and anthropometrics (BMI and height), while 16p11.2p exhibited significant associations with anthropometrics, schizophrenia, bipolar disorder, and Alzheimer’s disease. Furthermore, 7q11.23, 3q29, and 22q11.2 each displayed significant associations with two phenotypes within the blood biomarkers category.

Height and HDL displayed the most associations with the genomic regions, totalling six and five associations, respectively. Seven phenotypes (PTSD, MDD, ADHD, cannabis use disorder, lung cancer, chronic kidney disease, and coronary artery disease) did not yield significant results with any of the regions. Analyzing phenotype category associations revealed that anthropometrics and blood biomarkers exhibited the highest number of significant associations (8). Only one association was significant for the substance use category: 16p11.2d with the AUDIT score. Similarly, the association between 16p11.2p and T2D was the only significant association in the somatic disorders category.

There was no significant correlation between region size and the number of associations with traits (r_s_=-0.18, p-value=0.67), indicating that larger regions do not necessarily exhibit a greater number of associations due to an increased base pairs count within those regions.

### Relation to genetic architecture

We further explored factors relevant to the number of associations for traits with regions, utilizing the MiXeR tool to quantify the traits’ polygenicity (Table S2, Figure S1). No significant correlation was found between polygenicity and Bonferroni-adjusted associations of regions with traits (r_s_=0.07, p=0.76). However, a significant correlation was observed between heritability and the number of associations (r_s_=0.55, p=0.01), suggesting that the traits may have a genetic architecture where a smaller number of genes with larger effects contribute to the heritability.

### Partitioned heritability via GSA-MiXeR

We delved deeper into the observed MAGMA phenotype-region associations by employing GSA-MiXeR, which accounts for region size and local genomic characteristics. The highest partitioned heritability estimate was found for 16p11.2p, explaining 2.43×10^−3^ h2 of WBC, for 1q21.1d explaining ASD (1.33×10^−3^), and 3q29 for WBC (1.02×10^−3^) (Figure 2A). The maximum partitioned heritability across all regions was estimated for WBC (4.26×10^−3^), ASD (2.92×10^−3^), height (2.55×10^−3^), and schizophrenia (2.12×10^−3^). There was no significant difference in partitioned heritability estimates between GSA-MiXeR and LDAK-GBAT (t=-0.60, p=0.55) (Table S3, Figure S2-S3).

**Figure 2.**
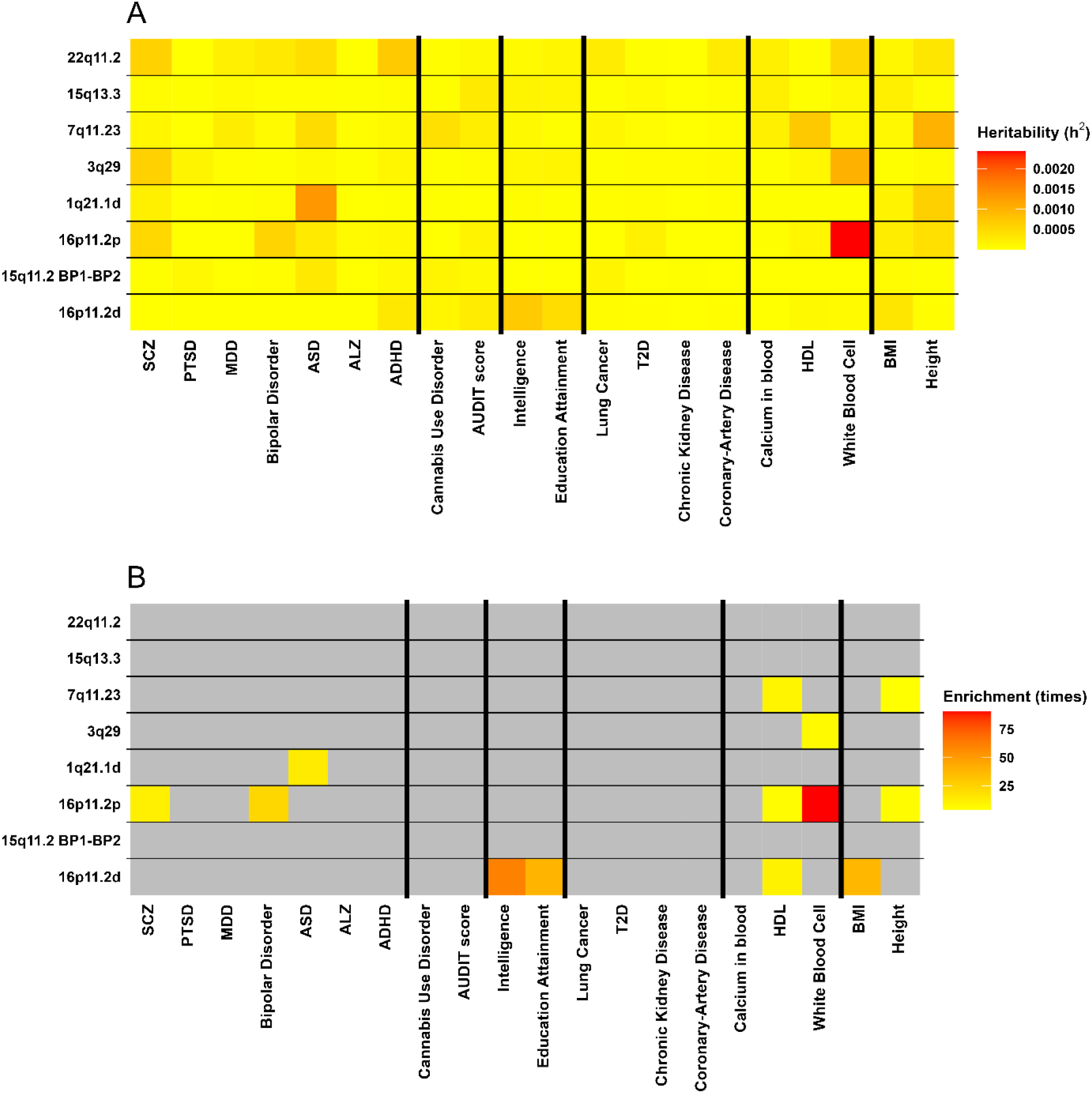
Estimates of SNP heritability (A) and fold enrichment estimates (in times) (B) for 8 regions of interest for 20 traits by GSA-MiXeR. Regions of interest are sorted in decreasing order on the x-axis, and traits are categorized on the y-axis. Colour denotes enriched regions for traits. ADHD = Attention Deficit Hyperactivity Disorder, AUDIT score = Alcohol Use Disorders Identification Test (AUDIT) score, ALZ = Alzheimer’s disease, ASD = Autism Spectrum Disorder, BMI = Body Mass Index, HDL = High-Density Lipoprotein, MDD = Major Depressive Disorder, PTSD = Post-Traumatic Stress Disorder, SCZ = Schizophrenia, T2D = Type II Diabetes.

### Enrichment

While MAGMA identifies genomic regions significantly associated with a phenotype, it lacks quantitative measures of association magnitude, while GSA-MiXeR enrichment quantifies fold enrichment, providing insight into the strength of these associations. Following correction for SD, 8% (13) of the region-phenotype pairs were enriched. Maximum enrichments were observed for the association between 16p11.2p and WBC (90.57), 16p11.2d and intelligence (60.98), AUDIT score (58.97), educational attainment (38.75), and BMI (37.54) (Figure 2B). The most enriched regions were 16p11.2p (5) and 16p11.2d (4), with no correlation between the number of enriched associations and region size (r_s_=-0.50, p=0.20).

Given the distinct computational parameters of MAGMA and GSA-MiXeR, we aimed to compare the outcomes generated by both tools. Genomic regions with significant p-values in MAGMA, coupled to pronounced enrichment in GSA-MiXeR, provide compelling evidence for their association with specific phenotypes. Out of 27 region-phenotype pairs significant in MAGMA across all investigated regions, 12 pairs were significantly enriched. The association between 16p11.2p and WBC exhibited the strongest enrichment, despite not being significant in MAGMA. The correlation between the number of MAGMA-significant phenotype-region associations and enriched region-phenotype pairs was significant (r_s_=0.75, p=0.03).

Traits with the highest number of enrichments in the regions of interest were HDL (3), WBC (2), and height (2), whereas traits with the highest number of associations in MAGMA gene-based analysis were height (6), HDL (5), and schizophrenia (3). Overall, both MAGMA and GSA-MiXeR revealed significant associations and enrichments for anthropometrics, blood biomarkers, and mental disorders (Table S4).

We evaluated the collective enrichment of all regions. Our goal was to determine if CNV-containing regions have a greater impact on specific phenotypes compared to other genomic regions of similar size. We found enrichment for 6 phenotypes, with the highest observed for WBC (5.34), PTSD (4.21), and AUDIT score (4.05) (Table S5).

### Gene-based for the genes in the regions of interest

We conducted a gene-level analysis within the genomic regions of interest using MAGMA (Figure S4). The list of genes in these regions, according to NCBI 37.3 gene definitions, can be found in Table S6.

The most frequently significant gene was *DOC2A*, associated with 11 traits. Following closely, *ATP2A1, ATXN2L, MLXIPL, RABEP2, SH2B1, TAOK2, TBX6, TMEM21*9, and *TUFM* were significant for 10 traits each. *ASPHD1, CD19, COMT, INO80E, KCTD13, SPNS1, TBL2*, and *TLCD3B* were significant for 9 traits each (Figure S4). Notably, 8 genes belonged to the 16p11.2p region, and 7 genes to the 16p11.2d region, which had the highest number of significant associations and enrichments with traits.

Twenty genes were significant for only one trait, with 7 of them located in the 22q11.2 region, the largest region investigated. None of these genes belonged to the smallest region, 16p11.2d. The correlation between the number of significant gene-trait associations in a region and the size of the region was not significant (r_s_=0.12, p=0.79).

To determine whether associations between the region and phenotype are evenly distributed across its genes or driven by specific genes, we compared the significance of the entire region to that of individual genes within it. Figure 3 depicts this comparison using -log10 p-values from MAGMA analysis for SCZ and HDL, data for other traits are in Figure S5. The findings reveal that the entire 16p11.2p region for SCZ and HDL exhibited values exceeding 1.96×SD from the mean of individual genes results within the region. However, for the 15q13.3 region, varying significance levels among separate genes versus the whole region suggest that some genes cancel out others.

**Figure 3.**
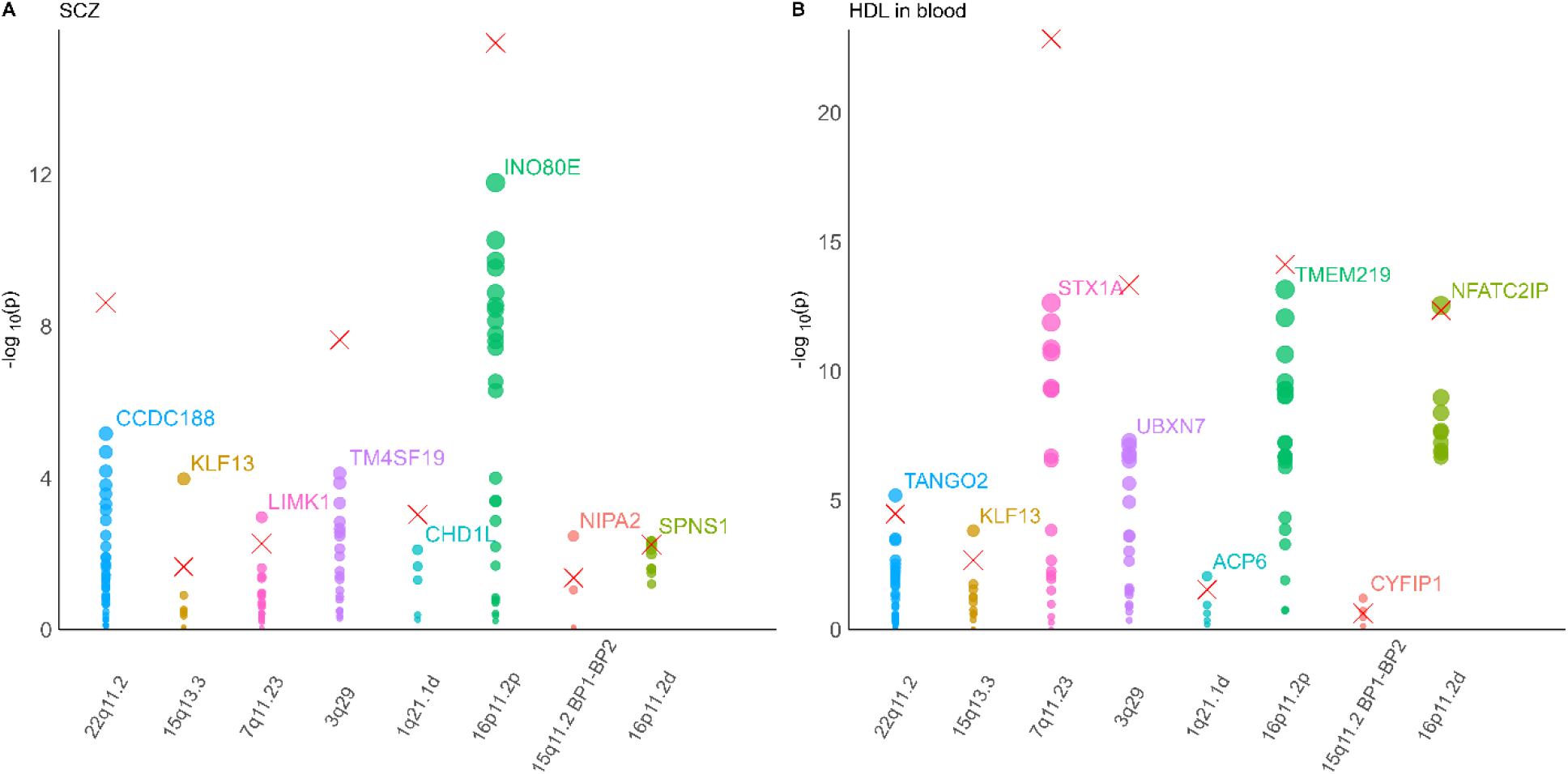
-log10 (p-values) for the individual genes in regions and the whole region of interest (cross) in MAGMA gene analysis for schizophrenia (SCZ) (A) and high density lipoprotein (HDL) levels in blood (B). The top gene is labelled.

In all traits, 35 gene associations (22%) exceeded the 1.96xSD threshold from the mean of associations for region-specific genes. Schizophrenia (5), HDL (5), educational attainment (4), and intelligence (4) exhibited the highest counts of significant associations for the entire region, implying a dependence on coordinated gene action within biological pathways rather than individual genes.

## Discussion

CNVs play important roles in disease and evolution, yet their links with clinical and functional traits present challenges due to diverse phenotypic manifestations and the influence of other genetic and environmental factors on CNV penetrance (1,48). Limited data from CNV carriers hinders research progress. To address this, we investigated the relationship between eight pathogenic CNVs and 20 complex traits by integrating common genetic variation within the CNV-containing genomic regions.

Most significant associations with traits and disorders identified by MAGMA were observed in the 16p11.2d and 16p11.2p regions and our analysis confirmed that region size does not account for associations number. CNVs within the 16p11.2 region have been linked to various neurodevelopmental disorders (49), including ASD (50), schizophrenia (51), ADHD (52) and intellectual disability (53,54) as well as obesity (11), cardiovascular conditions (13) and psychiatric disorders (55). However, only specific associations for this region, such as with schizophrenia, cognition, and anthropometrics, reached significance in our study. Our findings align with a previous study, which highlighted the 16p11.2 locus’s associations with blood biomarkers and anthropometric variables (12). Notably, associations with Alzheimer’s disease, AUDIT score, T2D, and HDL, uncommon in the literature, lacked clear clinical representation in CNV-related syndromes for this region. Additionally, these traits exhibited lower polygenicity compared to others in our analysis. Unusual associations could stem from non-coding variants influencing gene regulation, context-dependent factors (e.g., tissue specificity, developmental stage), or unveil novel biological pathways or functions.

The number of associations with regions showed no significant correlation with the polygenicity of traits and disorders. However, a strong correlation between the number of associations and heritability suggests that the increased associations between CNVs and specific traits arise from a greater genetic influence on phenotypic variability, rather than environmental factors. This may be attributed to distinct chromatin accessibility in the 16p11.2d and 16p11.2p regions compared to non-CNV regions, e.g. shown in human brain tissue (56). This differential accessibility may indicate varied availability for regulatory elements like transcription factors and microRNAs, impacting gene expression and function. Consequently, common genetic variants in these regions may exert regulatory effects on multiple traits, leading to significant associations and enrichment. Looking closer at individual genes in the CNV regions, we observed two distinct patterns: 1) when the overall association of the region with the phenotype surpassed that of specific genes within it; 2) when particular genes or clusters of genes exhibited notably stronger associations with the phenotype compared to the CNV region as a whole. Even though, CNV regions contain many genes that are strongly related to certain phenotypes, non-enriched genes in these regions could dilute the association and enrichment of the whole region. Among the specific genes in these regions, the genes that were associated with most of the phenotypes were located in 16p11.2p and 16p11.2d, which might also explain why they have the greatest number of significant associations. Moreover, two genes (*MLXIPL* and *TBL2*) from the 7q11.23 region, along with the *COMT* gene from the 22q11.2 region, exhibited significant associations with over 10 traits. *COMT*, a well-studied gene in the 22q11.2 region, is implicated in cognitive and psychiatric features in individuals with 22q11.2 deletion syndrome, including the risk of schizophrenia-like symptoms (57,58). *TBL2* is linked to Williams-Beuren syndrome, while *MLXIPL* is associated with impaired glucose tolerance and diabetes mellitus in carriers of the 7q11.23 CNV (59)

The gene with the highest number of trait associations, *DOC2A*, plays a role in synaptic transmission and calcium-dependent exocytosis regulation (60). Functionally, five genes are linked to ion transport and signalling (61–65), five to transcriptional regulation and development (66–70), two to protein translation and metabolism (71,72), *COMT* to enzymatic activity (73), *CD19* to immune system regulation (74), *ATXN2L* to RNA metabolism (75) and functions of two genes remain unclear. These genes collectively influence diverse phenotypes and traits, playing crucial roles in cellular processes. While many of these genes have been extensively studied in the context of CNV-related syndromes, not all have received adequate attention and may represent promising candidates for future research (49,76,77).

Genes that show the significance of a single phenotype is of particular interest. Especially, when the overall association of the genomic region is statistically significant, like for *SPN* for 16p11.2p or *GJA8* for 1q21.1d distal. The majority of these distinct genes exhibit lower significance than the overall region and are significant in fewer associations with phenotypes than other genes in the region. This suggests their specificity to the phenotypes, although their individual impact may not be substantial enough to drive changes in the overall region’s association with these phenotypes. In an alternative scenario, a unique gene may demonstrate significance while the entire genomic region does not, like *NIPA1* for 15q11.2 BP1-BP2 or *THAP7* for 22q11.2. This implies that not only is the individual gene insufficient to drive phenotypic changes, but there is also a lack of other statistically significant genes in the region to contribute to its overall influence. A possible theory is that the CNV regions might be enriched for genes that are involved in complex networks that influence complex traits or involve groups of genes that tend to be co-expressed or function together (12,78,79). Therefore, the common genetic variants in these regions may have synergistic or antagonistic effects on multiple traits, resulting in significant associations and enrichment.

We compared MAGMA gene-based analysis with another tool for assessing heritability and enrichment of specific CNVs. Both GSA-MiXeR and MAGMA identified significant associations and enrichments primarily in the 16p11.2d and 16p11.2p regions, thereby validating each other’s findings.

Maximum enrichments were observed for WBC, intelligence, and educational attainment, underscoring the need for a detailed investigation of these traits in CNV carriers. Notably, WBC exhibited over 90 times enrichment, warranting particular attention. Additionally, height score and HDL displayed notable enrichments across all regions. Traits associated with anthropometrics, blood markers, and mental disorders showed the highest number of enrichments with regions, while substance use and somatic disorders lacked enrichment. Clinical heterogeneity may influence the relationship between phenotype categories and enrichments, potentially due to varying effect sizes of variants across different disorder subtypes. WBC, influenced by diverse factors including genetics, emphasizes the possibility of region-specific genetic contributions. Although MAGMA analysis did not yield significance for the WBC-16p11.2p pair despite its high enrichment score, GSA-MiXeR may offer novel gene candidates for further exploration, based on complementary MAGMA analysis and biologically relevant gene sets, as recommended by the authors (46).

The enrichments for all the CNVs-phenotype associations were lower than for particular CNV-phenotype associations leading to the conclusion that they should be considered separately.

Our study’s scope was limited to eight CNV regions linked to neurodevelopmental disorders, which may affect the breadth of our conclusions. We analyzed 20 traits and disorders across six categories with uneven phenotype distribution, complicating cross-category analysis. The GWAS quality and size per phenotype could impact our findings, though we initially screened out traits with inadequate GWAS data. We advise using MAGMA and GSA-MiXeR together for a comprehensive interpretation — MAGMA for statistical significance and GSA-MiXeR for region’s enrichment. It’s important to note that common variation may serve as a proxy for just a portion of CNV effects, and these are influenced by other genetic and environmental factors.

Our study could be enhanced by including a broader range of CNV regions and phenotypes relevant to human health. Further analysis could investigate the effects of CNV segments, not only on individual genes but also on gene groups or combinations across different CNV regions. Tools like LDAK-GBAT, used in conjunction with GSA-MiXeR, could add validation and replication to our results. Since CNV deletions and duplications can influence gene expression nearby, examining expression quantitative trait loci (eQTL) within these regions could elucidate the molecular mechanisms behind our observed associations.

This approach also allows for the exploration of new genes within CNV regions and the investigation of less-studied phenotypes, such as neurodegenerative disorders. By utilizing publicly available data from diverse populations, we can overcome the limitations of scarce patient cohort data. Determining the role of individual genes in these chromosomal regions could lead to the discovery of new genes for molecular experimentation.

## Conclusion

We demonstrated the versatility of the current advanced tools for gene-set analysis, which can be applied not just to genes but also to broader genomic regions, including those that are susceptibility areas for CNVs.

## Data Availability

All data produced in the present study are available upon reasonable request to the authors

## References

1. Pös O, Radvanszky J, Buglyó G, Pös Z, Rusnakova D, Nagy B, Szemes T (2021): DNA copy number variation: Main characteristics, evolutionary significance, and pathological aspects. Biomed J 44: 548–559.

2. Redon R, Ishikawa S, Fitch KR, Feuk L, Perry GH, Andrews TD, et al. (2006): Global variation in copy number in the human genome. Nature 2006 444:7118 444: 444–454.

3. Sebat J, Lakshmi B, Troge J, Alexander J, Young J, Lundin P, et al. (2004): Large-scale copy number polymorphism in the human genome. Science 305: 525–528.

4. Itsara A, Cooper GM, Baker C, Girirajan S, Li J, Absher D, et al. (2009): Population analysis of large copy number variants and hotspots of human genetic disease. Am J Hum Genet 84: 148–161.

5. Grayton HHM, Fernandes C, Rujescu D, Collier DA (2012): Copy number variations in neurodevelopmental disorders. Prog Neurobiol 99: 81–91.

6. Rice AM, McLysaght A (2017): Dosage sensitivity is a major determinant of human copy number variant pathogenicity. Nature Communications 2017 8:1 8: 1–11.

7. Stranger BE, Forrest MS, Dunning M, Ingle CE, Beazlsy C, Thorne N, et al. (2007): Relative impact of nucleotide and copy number variation on gene expression phenotypes. Science 315: 848.

8. Lupski JR (2007): Genomic rearrangements and sporadic disease. Nature Genetics 2007 39:7 39: S43–S47.

9. Rees E, Kirov G (2021): Copy number variation and neuropsychiatric illness. Curr Opin Genet Dev 68: 57–63.

10. Kendall KM, Bracher-Smith M, Fitzpatrick H, Lynham A, Rees E, Escott-Price V, et al. (2019): Cognitive performance and functional outcomes of carriers of pathogenic copy number variants: analysis of the UK Biobank. The British Journal of Psychiatry 214: 297–304.

11. Macé A, Tuke MA, Deelen P, Kristiansson K, Mattsson H, Nõukas M, et al. (2017): CNV-association meta-analysis in 191,161 European adults reveals new loci associated with anthropometric traits. Nature Communications 2017 8:1 8: 1–11.

12. Auwerx C, Lepamets M, Sadler MC, Patxot M, Stojanov M, Baud D, et al. (2022): The individual and global impact of copy-number variants on complex human traits. Am J Hum Genet 109.

13. Costain G, Silversides CK, Bassett AS (2016): The importance of copy number variation in congenital heart disease. npj Genomic Medicine 2016 1:1 1: 1–11.

14. Shlien A, Malkin D (2009): Copy number variations and cancer. Genome Med 1: 1–9.

15. Crawford K, Bracher-Smith M, Owen D, Kendall KM, Rees E, Pardiñas AF, et al. (2019): Medical consequences of pathogenic CNVs in adults: analysis of the UK Biobank. J Med Genet 56: 131–138.

16. Pös O, Radvanszky J, Styk J, Pös Z, Buglyó G, Kajsik M, et al. (2021): Copy Number Variation: Methods and Clinical Applications. Applied Sciences 11. 10.3390/app11020819

17. Li W, Olivier M (2013): Current analysis platforms and methods for detecting copy number variation. Physiol Genomics 45: 1–16.

18. Craddock N, Hurles ME, Cardin N, Pearson RD, Plagnol V, Robson S, et al. (2010): Genome-wide association study of copy number variation in 16,000 cases of eight common diseases and 3,000 shared controls. Nature 464: 713.

19. Uffelmann E, Huang QQ, Munung NS, de Vries J, Okada Y, Martin AR, et al. (2021): Genome-wide association studies. Nature Reviews Methods Primers 2021 1:1 1: 1–21.

20. Kirov G, Rees E, Walters JTRR, Escott-Price V, Georgieva L, Richards AL, et al. (2014): The penetrance of copy number variations for schizophrenia and developmental delay. Biol Psychiatry 75: 378–385.

21. Ehrhart F, Silva A, Amelsvoort T van, Scheibler E von, Evelo C, Linden DEJ (2022): Converging pathways found in copy number variation syndromes with high schizophrenia risk. bioRxiv 2022.02.07.479370.

22. Vaez M, Montalbano S, Sánchez XC, Hellberg K-LG, Dehkordi SR, Krebs MD, et al. (2023): Population-based Risk of Psychiatric Disorders Associated with Recurrent CNVs. medRxiv 2023.09.04.23294975.

23. Owen D, Bracher-Smith M, Kendall KM, Rees E, Einon M, Escott-Price V, et al. (2018): Effects of pathogenic CNVs on physical traits in participants of the UK Biobank. BMC Genomics 19: 1–9.

24. Demontis D, Walters RK, Martin J, Mattheisen M, Als TD, Agerbo E, et al. (2018): Discovery of the first genome-wide significant risk loci for attention deficit/hyperactivity disorder. Nature Genetics 2018 51:1 51: 63–75.

25. Jansen IE, Savage JE, Watanabe K, Bryois J, Williams DM, Steinberg S, et al. (2019): Genome-wide meta-analysis identifies new loci and functional pathways influencing Alzheimer’s disease risk. Nat Genet 51: 404–413.

26. Grove J, Ripke S, Als TD, Mattheisen M, Walters RK, Won H, et al. (2019): Identification of common genetic risk variants for autism spectrum disorder. Nat Genet 51: 431–444.

27. Mullins N, Forstner AJ, O’Connell KS, Coombes B, Coleman JRI, Qiao Z, et al. (2021): Genome-wide association study of more than 40,000 bipolar disorder cases provides new insights into the underlying biology. Nat Genet 53: 817–829.

28. Wray NR, Ripke S, Mattheisen M, Trzaskowski M, Byrne EM, Abdellaoui A, et al. (2018): Genome-wide association analyses identify 44 risk variants and refine the genetic architecture of major depression. Nature Genetics 2018 50:5 50: 668–681.

29. Nievergelt CM, Maihofer AX, Klengel T, Atkinson EG, Chen CY, Choi KW, et al. (2019): International meta-analysis of PTSD genome-wide association studies identifies sex- and ancestry-specific genetic risk loci. Nature Communications 2019 10:1 10: 1–16.

30. Trubetskoy V, Pardiñas AF, Qi T, Panagiotaropoulou G, Awasthi S, Bigdeli TB, et al. (2022): Mapping genomic loci implicates genes and synaptic biology in schizophrenia. Nature 2022 604:7906 604: 502–508.

31. Sanchez-Roige S, Palmer AA, Fontanillas P, Elson SL, Adams MJ, Howard DM, et al. (2019): Genome-wide association study meta-analysis of the alcohol use disorders identification test (AUDIT) in two population-based cohorts. American Journal of Psychiatry 176: 107–118.

32. Johnson EC, Demontis D, Thorgeirsson TE, Walters RK, Polimanti R, Hatoum AS, et al. (2020): A large-scale genome-wide association study meta-analysis of cannabis use disorder. Lancet Psychiatry 7: 1032–1045.

33. Lee JJ, Wedow R, Okbay A, Kong E, Maghzian O, Zacher M, et al. (2018): Gene discovery and polygenic prediction from a genome-wide association study of educational attainment in 1.1 million individuals. Nature Genetics 2018 50:8 50: 1112–1121.

34. Savage JE, Jansen PR, Stringer S, Watanabe K, Bryois J, De Leeuw CA, et al. (2018): Genome-wide association meta-analysis in 269,867 individuals identifies new genetic and functional links to intelligence. Nature Genetics 2018 50:7 50: 912–919.

35. Nikpay M, Goel A, Won HH, Hall LM, Willenborg C, Kanoni S, et al. (2015): A comprehensive 1000 Genomes–based genome-wide association meta-analysis of coronary artery disease. Nature Genetics 2015 47:10 47: 1121–1130.

36. Wuttke M, Li Y, Li M, Sieber KB, Feitosa MF, Gorski M, et al. (2019): A catalog of genetic loci associated with kidney function from analyses of a million individuals. Nature Genetics 2019 51:6 51: 957–972.

37. Mahajan A, Wessel J, Willems SM, Zhao W, Robertson NR, Chu AY, et al. (2018): Refining the accuracy of validated target identification through coding variant fine-mapping in type 2 diabetes. Nature Genetics 2018 50:4 50: 559–571.

38. Byun J, Han Y, Li Y, Xia J, Long E, Choi J, et al. (2022): Cross-ancestry genome-wide meta-analysis of 61,047 cases and 947,237 controls identifies new susceptibility loci contributing to lung cancer. Nat Genet 54: 1167–1177.

39. Astle WJ, Elding H, Jiang T, Allen D, Ruklisa D, Mann AL, et al. (2016): The Allelic Landscape of Human Blood Cell Trait Variation and Links to Common Complex Disease. Cell 167: 1415-1429.e19.

40. Graham SE, Clarke SL, Wu KHH, Kanoni S, Zajac GJM, Ramdas S, et al. (2021): The power of genetic diversity in genome-wide association studies of lipids. Nature 2021 600:7890 600: 675–679.

41. Sinnott-Armstrong N, Tanigawa Y, Amar D, Mars N, Benner C, Aguirre M, et al. (2021): Genetics of 35 blood and urine biomarkers in the UK Biobank. Nature Genetics 2021 53:2 53: 185–194.

42. Locke AE, Kahali B, Berndt SI, Justice AE, Pers TH, Day FR, et al. (2015): Genetic studies of body mass index yield new insights for obesity biology. Nature 2015 518:7538 518: 197–206.

43. de Leeuw CA, Mooij JM, Heskes T, Posthuma D (2015): MAGMA: Generalized Gene-Set Analysis of GWAS Data. PLoS Comput Biol 11: e1004219.

44. Holland D, Frei O, Desikan R, Fan CC, Shadrin AA, Smeland OB, et al. (2020): Beyond SNP heritability: Polygenicity and discoverability of phenotypes estimated with a univariate Gaussian mixture model. PLoS Genet 16: e1008612.

45. Frei O, Holland D, Smeland OB, Shadrin AA, Fan CC, Maeland S, et al. (2019): Bivariate causal mixture model quantifies polygenic overlap between complex traits beyond genetic correlation. Nature Communications 2019 10:1 10: 1–11.

46. Frei O, Hindley G, Shadrin AA, van der Meer D, Akdeniz BC, Hagen E, et al. (2024): Improved functional mapping of complex trait heritability with GSA-MiXeR implicates biologically specific gene sets. Nature Genetics 2024 56:6 56: 1310–1318.

47. Berrandou TE, Balding D, Speed D (2023): LDAK-GBAT: Fast and powerful gene-based association testing using summary statistics. The American Journal of Human Genetics 110: 23–29.

48. Zhang F, Gu W, Hurles ME, Lupski JR (2009): Copy Number Variation in Human Health, Disease, and Evolution. Annu Rev Genomics Hum Genet 10: 451–481.

49. Rein B, Yan Z (2020): 16p11.2 copy number variations and neurodevelopmental disorders. Trends Neurosci 43: 886–901.

50. Weiss LA, Shen Y, Korn JM, Arking DE, Miller DT, Fossdal R, et al. (2008): Association between microdeletion and microduplication at 16p11.2 and autism. N Engl J Med 358: 667–675.

51. Marshall CR, Howrigan DP, Merico D, Thiruvahindrapuram B, Wu W, Greer DS, et al. (2017): Contribution of copy number variants to schizophrenia from a genome-wide study of 41,321 subjects. Nat Genet 49: 27–35.

52. Gudmundsson OO, Walters GB, Ingason A, Johansson S, Zayats T, Athanasiu L, et al. (2019): Attention-deficit hyperactivity disorder shares copy number variant risk with schizophrenia and autism spectrum disorder. Transl Psychiatry 9: 258.

53. Hanson E, Nasir RH, Fong A, Lian A, Hundley R, Shen Y, et al. (2010): Cognitive and behavioral characterization of 16p11.2 deletion syndrome. J Dev Behav Pediatr 31: 649–657.

54. D’Angelo D, Lebon S, Chen Q, Martin-Brevet S, Snyder LAG, Hippolyte L, et al. (2016): Defining the Effect of the 16p11.2 Duplication on Cognition, Behavior, and Medical Comorbidities. JAMA Psychiatry 73: 20.

55. Niarchou M, Chawner SJRA, Doherty JL, Maillard AM, Jacquemont S, Chung WK, et al. (2019): Psychiatric disorders in children with 16p11.2 deletion and duplication. Transl Psychiatry 9. 10.1038/S41398-018-0339-8

56. Loviglio MN, Leleu M, Männik K, Passeggeri M, Giannuzzi G, van der Werf I, et al. (2017): Chromosomal contacts connect loci associated with autism, BMI and head circumference phenotypes. Mol Psychiatry 22: 836.

57. Karayiorgou M, Gogos JA (2004): The molecular genetics of the 22q11-associated schizophrenia. Molecular Brain Research 132: 95–104.

58. Gothelf D, Eliez S, Thompson T, Hinard C, Penniman L, Feinstein C, et al. (2005): COMT genotype predicts longitudinal cognitive decline and psychosis in 22q11.2 deletion syndrome. Nature Neuroscience 2005 8:11 8: 1500–1502.

59. Merla G, Brunetti-Pierri N, Micale L, Fusco C (2010): Copy number variants at Williams-Beuren syndrome 7q11.23 region. Hum Genet 128: 3–26.

60. Wang Q-W, Qin J, Chen Y-F, Tu Y, Xing Y-Y, Wang Y, et al. (2023): 16p11.2 CNV gene Doc2α functions in neurodevelopment and social behaviors through interaction with Secretagogin. Cell Rep 42: 112691.

61. Pan Y, Zvaritch E, Tupling AR, Rice WJ, De Leon S, Rudnicki M, et al. (2003): Targeted disruption of the ATP2A1 gene encoding the sarco(endo)plasmic reticulum Ca2+ ATPase isoform 1 (SERCA1) impairs diaphragm function and is lethal in neonatal mice. J Biol Chem 278: 13367–13375.

62. Doche ME, Bochukova EG, Su HW, Pearce LR, Keogh JM, Henning E, et al. (2012): Human SH2B1 mutations are associated with maladaptive behaviors and obesity. J Clin Invest 122: 4732–4736.

63. Yadav S, Oses-Prieto JA, Peters CJ, Zhou J, Pleasure SJ, Burlingame AL, et al. (2017): TAOK2 Kinase Mediates PSD95 Stability and Dendritic Spine Maturation through Septin7 Phosphorylation. Neuron 93: 379–393.

64. Kofler N, Corti F, Rivera-Molina F, Deng Y, Toomre D, Simons M (2018): The Rab-effector protein RABEP2 regulates endosomal trafficking to mediate vascular endothelial growth factor receptor-2 (VEGFR2)-dependent signaling. J Biol Chem 293: 4805–4817.

65. He M, Kuk ACY, Ding M, Chin CF, Galam DLA, Nah JM, et al. (2022): Spns1 is a lysophospholipid transporter mediating lysosomal phospholipid salvage. Proc Natl Acad Sci U S A 119. 10.1073/PNAS.2210353119

66. Abdul-Wahed A, Guilmeau S, Postic C (2017): Sweet Sixteenth for ChREBP: Established Roles and Future Goals. Cell Metab 26: 324–341.

67. Gao S, Li X, Amendt BA (2013): Understanding the Role of Tbx1 as a Candidate Gene for 22q11.2 Deletion Syndrome. Curr Allergy Asthma Rep 13: 613–621.

68. Gowans GJ, Schep AN, Wong KM, King DA, Greenleaf WJ, Morrison AJ (2018): INO80 Chromatin Remodeling Coordinates Metabolic Homeostasis with Cell Division. Cell Rep 22: 611–623.

69. Kizner V, Naujock M, Fischer S, Jäger S, Reich S, Schlotthauer I, et al. (2020): CRISPR/Cas9-mediated Knockout of the Neuropsychiatric Risk Gene KCTD13 Causes Developmental Deficits in Human Cortical Neurons Derived from Induced Pluripotent Stem Cells. Mol Neurobiol 57: 616–634.

70. Tsukumo Y, Tsukahara S, Furuno A, Iemura SI, Natsume T, Tomida A (2015): The endoplasmic reticulum-localized protein TBL2 interacts with the 60S ribosomal subunit. Biochem Biophys Res Commun 462: 383–388.

71. Choi CY, Vo MT, Nicholas J, Choi YB (2022): Autophagy-competent mitochondrial translation elongation factor TUFM inhibits caspase-8-mediated apoptosis. Cell Death Differ 29: 451–464.

72. Bertrand RE, Wang J, Xiong KH, Thangavel C, Qian X, Ba-Abbad R, et al. (2021): Ceramide synthase TLCD3B is a novel gene associated with human recessive retinal dystrophy. Genet Med 23: 488–497.

73. Craddock N, Owen MJ, O’Donovan MC (2006): The catechol-O-methyl transferase (COMT) gene as a candidate for psychiatric phenotypes: evidence and lessons. Mol Psychiatry 11: 446–458.

74. Wang K, Wei G, Liu D (2012): CD19: a biomarker for B cell development, lymphoma diagnosis and therapy. Exp Hematol Oncol 1: 36.

75. Ostrowski LA, Hall AC, Mekhail K (2017): Ataxin-2: From RNA Control to Human Health and Disease. Genes (Basel) 8: 2–21.

76. Du Q, de la Morena MT, van Oers NSC (2020): The Genetics and Epigenetics of 22q11.2 Deletion Syndrome. Front Genet 10: 486538.

77. Mervis CB, Morris CA, Klein-Tasman BP, Velleman SL, Osborne LR (2021): 7q11.23 Duplication Syndrome. GeneReviews®.

78. Xu Y, DuanMu H, Chang Z, Zhang S, Li Z, Li Z, et al. (2012): The application of gene co-expression network reconstruction based on CNVs and gene expression microarray data in breast cancer. Mol Biol Rep 39: 1627–1637.

79. Park C, Ahn J, Yoon Y, Park S (2012): Identification of functional CNV region networks using a CNV-gene mapping algorithm in a genome-wide scale. Bioinformatics 28: 2045–2051.

