## Supplementary Figures for "Association of complex traits with common genetic variation across genomic regions containing pathogenic copy number variations"

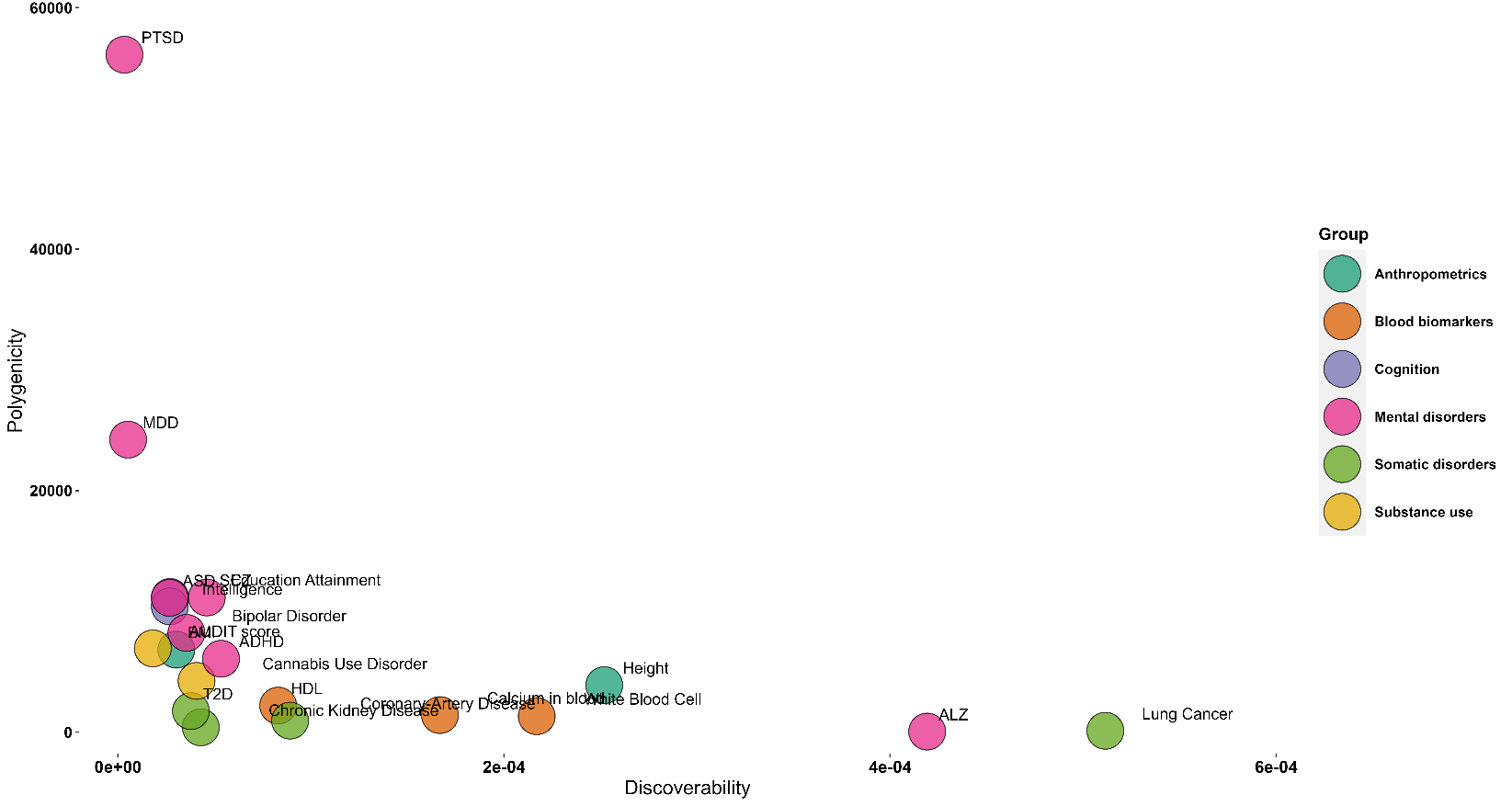

**Figure S1.** Polygenicity and discoverability of 20 traits. Polygenicity (proportion of causally associated SNPs) and discoverability (causal SNP effect size variance) were estimated using MiXeR. ADHD = Attention Deficit Hyperactivity Disorder, AUDIT score = Alcohol Use Disorders Identification Test (AUDIT) score, ALZ = Alzheimer’s disease, BMI = Body Mass Index, HDL = High-Density Lipoprotein, MDD = Major Depressive Disorder, PTSD = Post-Traumatic Stress Disorder, SCZ = schizophrenia, T2D = Type II Diabetes.

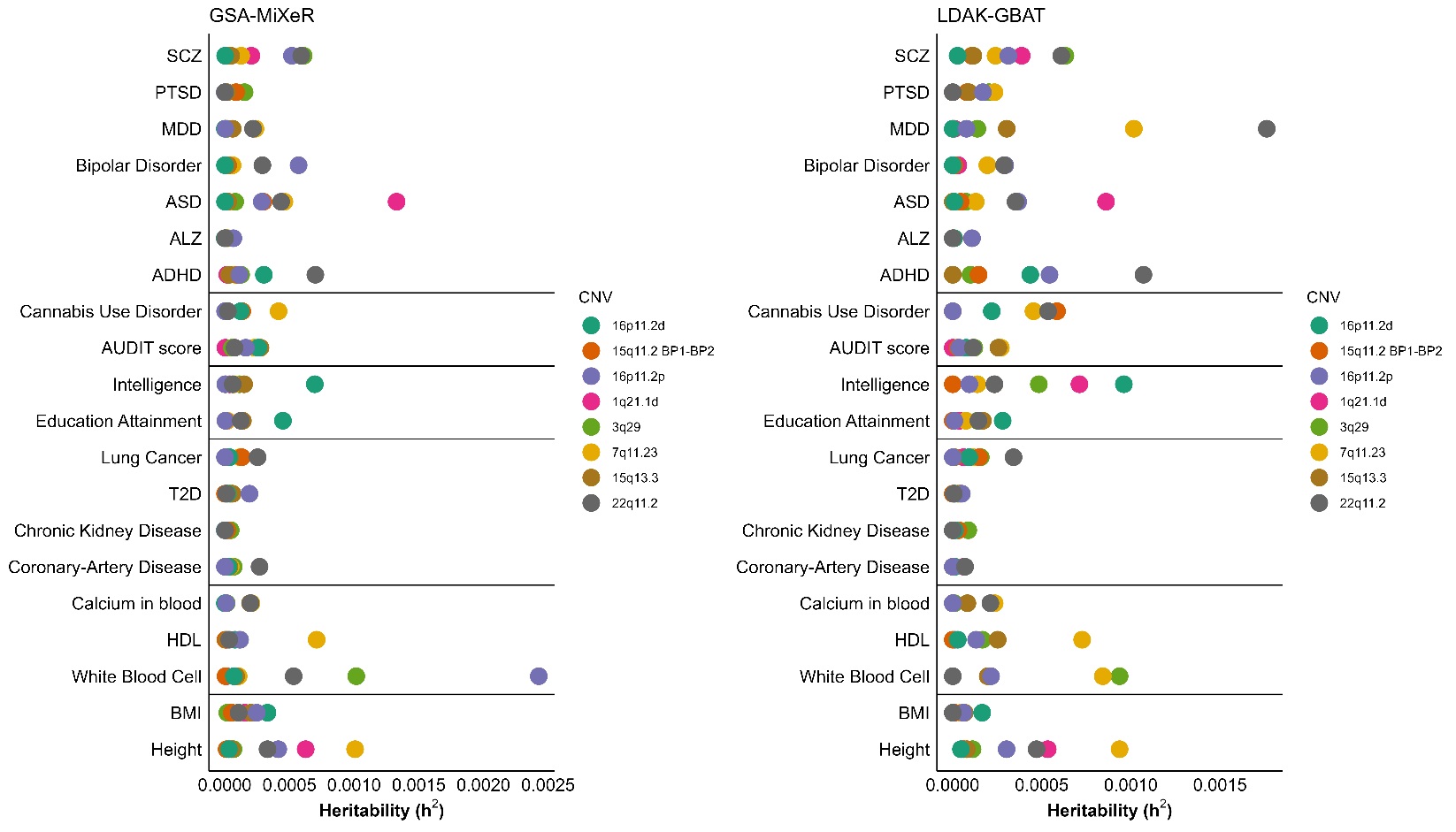

**Figure S2.** GSA-MiXeR heritability estimates (A) and LDAK-GBAT heritability estimates (B) for 20 traits and 8 genome regions of interest. The traits are categorized on the y-axis. ADHD = Attention Deficit Hyperactivity Disorder, AUDIT score = Alcohol Use Disorders Identification Test (AUDIT) score, ALZ = Alzheimer’s disease, ASD = Autism Spectrum Disorder, BMI = Body Mass Index, HDL = High-Density Lipoprotein, MDD = Major Depressive Disorder, PTSD = Post-Traumatic Stress Disorder, SCZ = Schizophrenia, T2D = Type II Diabetes.

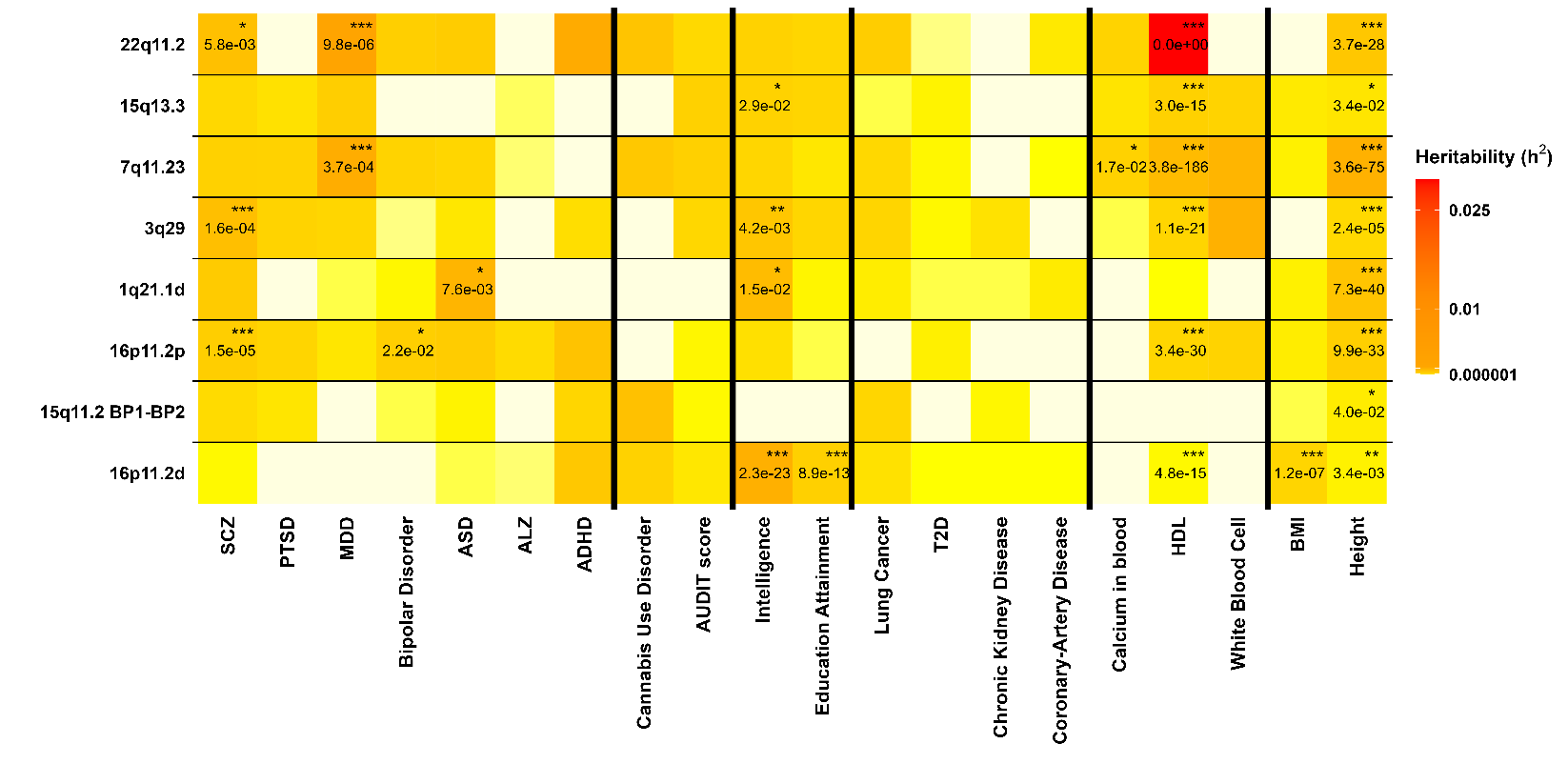
**Figure S3.** Estimates of SNP heritability for 8 regions of interest by via LDAK-GBAT for 20 traits. Regions of interest are sorted in decreasing order on the x-axis, and traits are categorized on the y-axis. The colour indicates the heritability with significance levels expressed by asterisks (*** for p-value ≤ 0.0005, ** for p-value ≤ 0.005, * for p-value ≤ 0.05). P-values equal to one are not represented. ADHD = Attention Deficit Hyperactivity Disorder, AUDIT score = Alcohol Use Disorders Identification Test (AUDIT) score, ALZ = Alzheimer’s disease, ASD = Autism Spectrum Disorder, BMI = Body Mass Index, HDL = High-Density Lipoprotein, MDD = Major Depressive Disorder, PTSD = Post-Traumatic Stress Disorder, SCZ = Schizophrenia, T2D = Type II Diabetes.

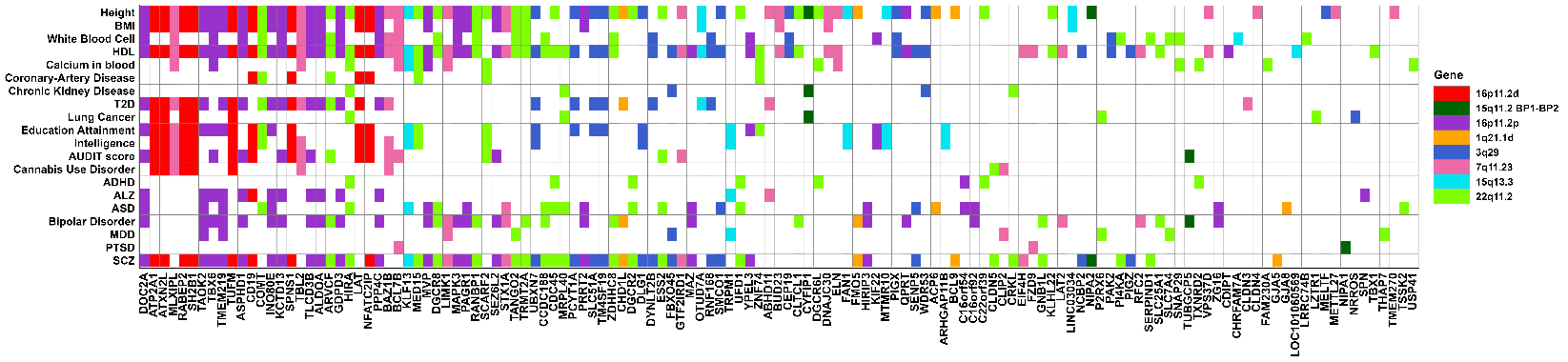

**Figure S4.** Genes in the CNV-containing regions that were significant in gene analysis using MAGMA. Genes are sorted in increasing order by number of significant associations on the x-axis, and traits are on the y-axis. Colour represents the CNV region to which gene belongs to. ADHD = Attention Deficit Hyperactivity Disorder, AUDIT score = Alcohol Use Disorders Identification Test (AUDIT) score, ALZ = Alzheimer’s disease, BMI = Body Mass Index, HDL = High-Density Lipoprotein, MDD = Major Depressive Disorder, PTSD = Post-Traumatic Stress Disorder, SCZ = schizophrenia, T2D = Type II Diabetes.

| 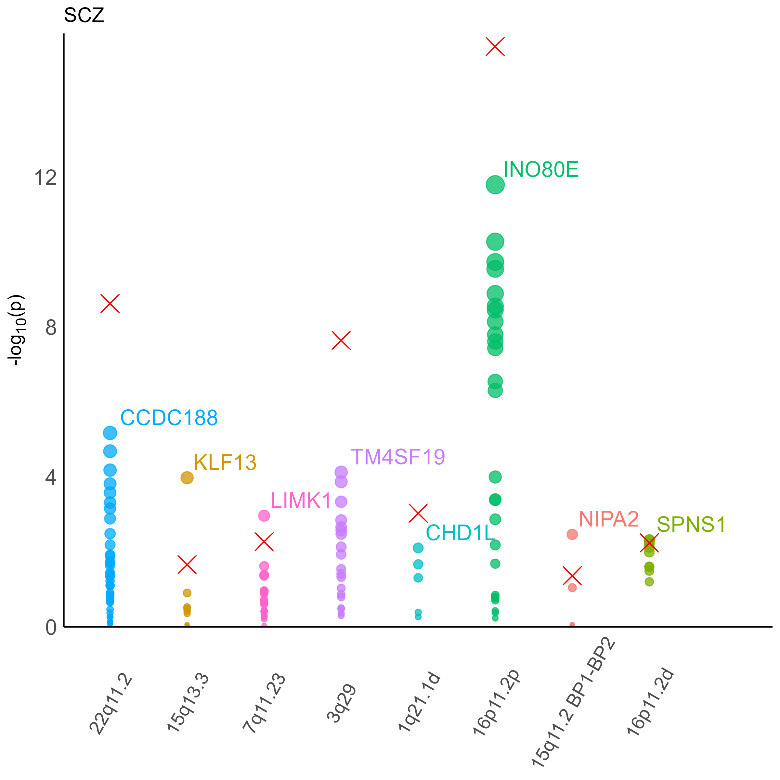 | 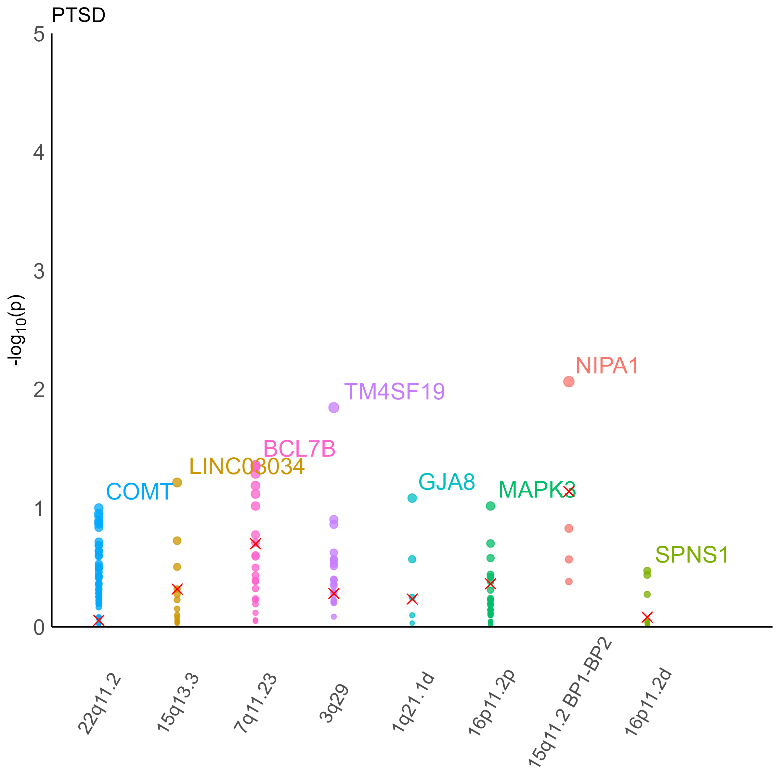 |
| --- | --- |
| 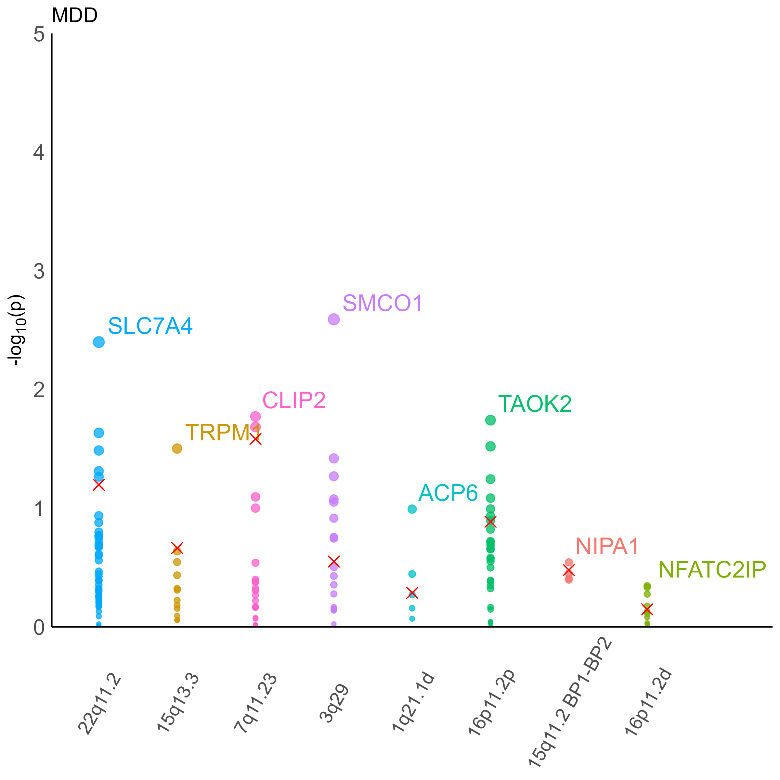 | 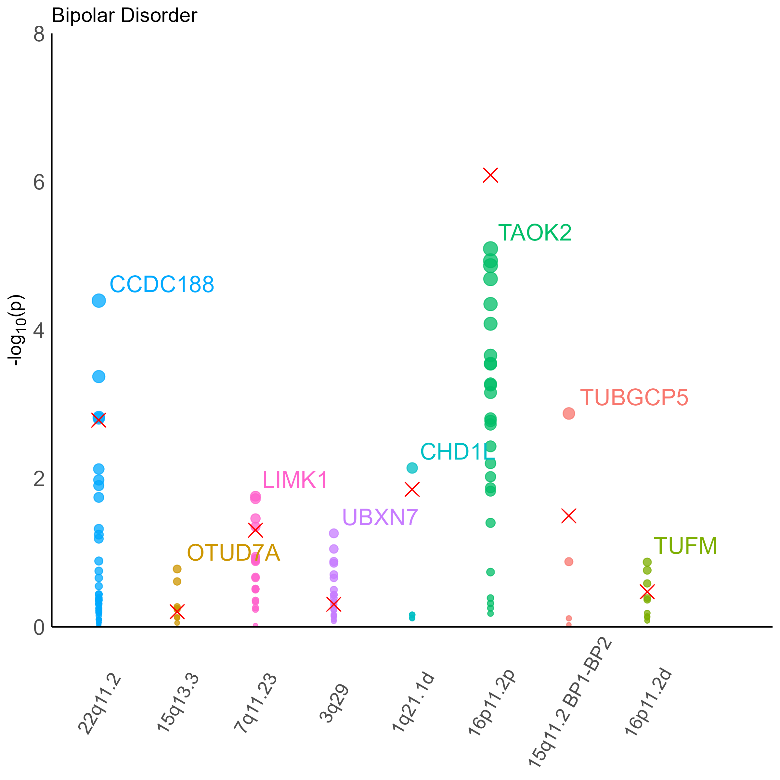 |
| 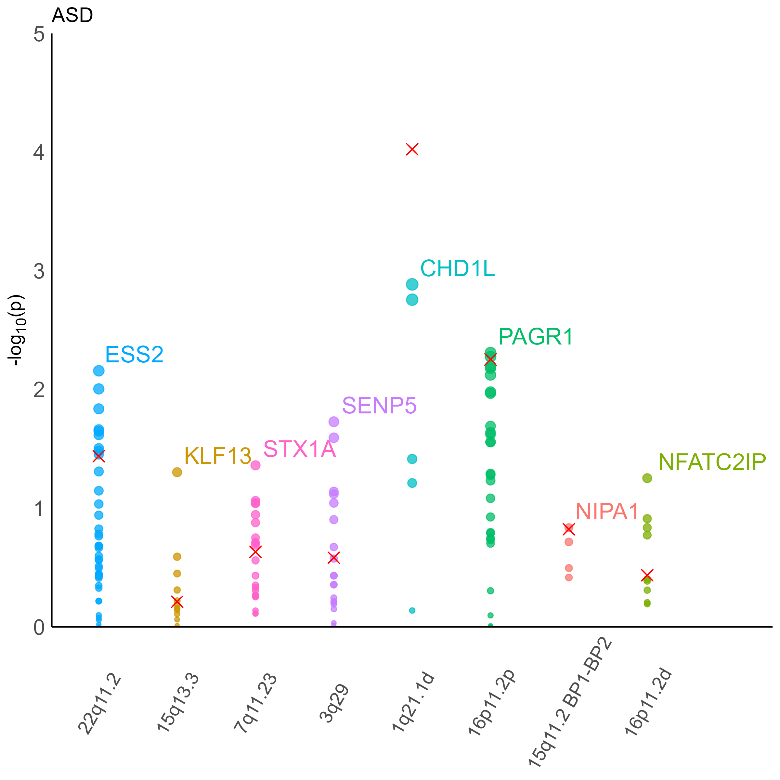 | 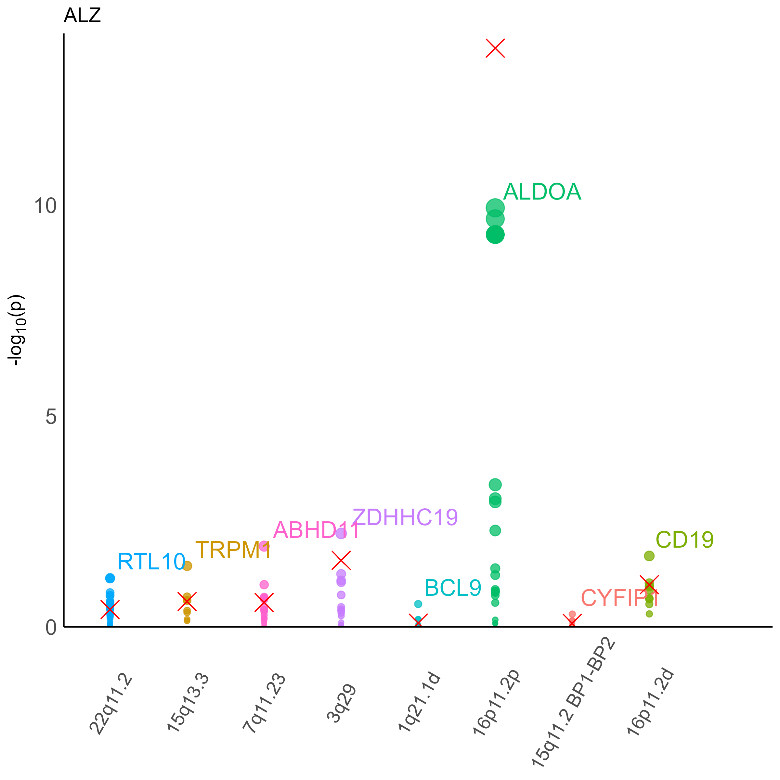 |
| 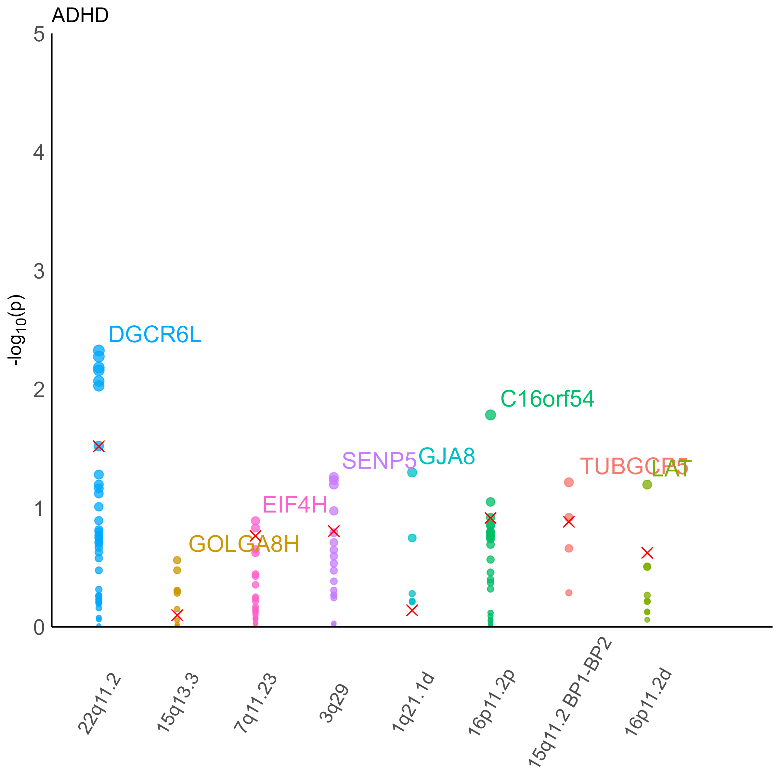 | 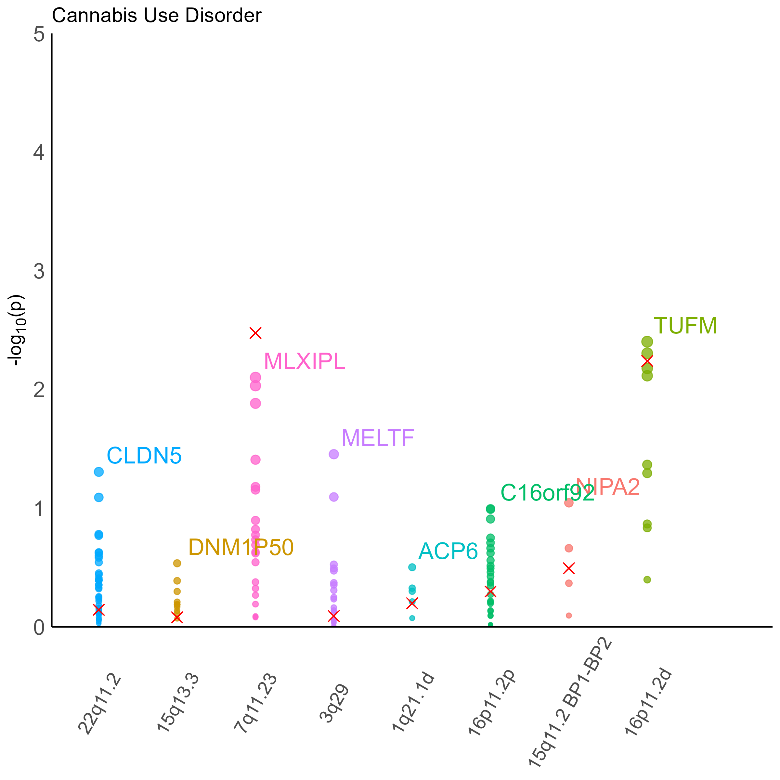 |
| 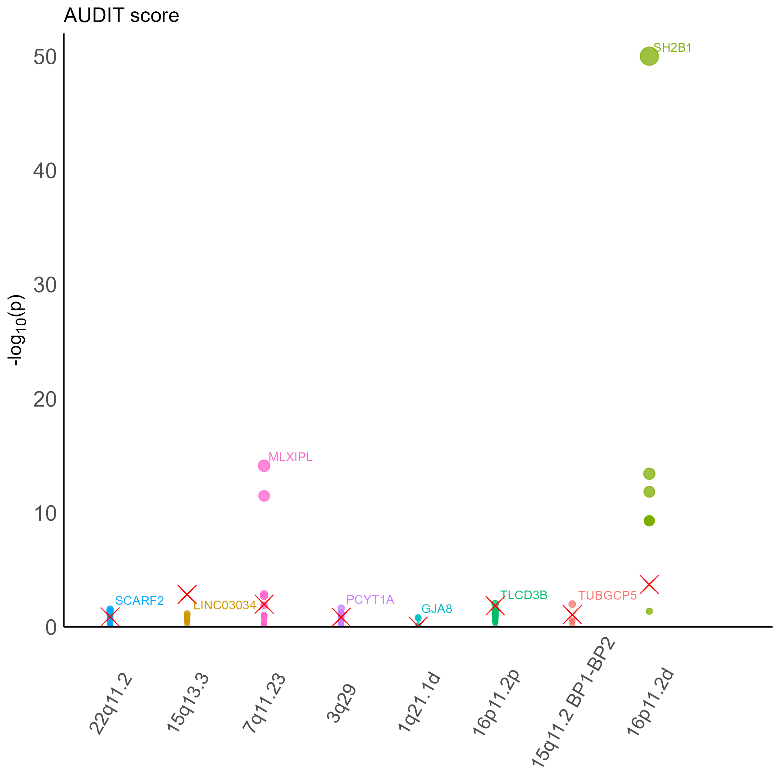 | 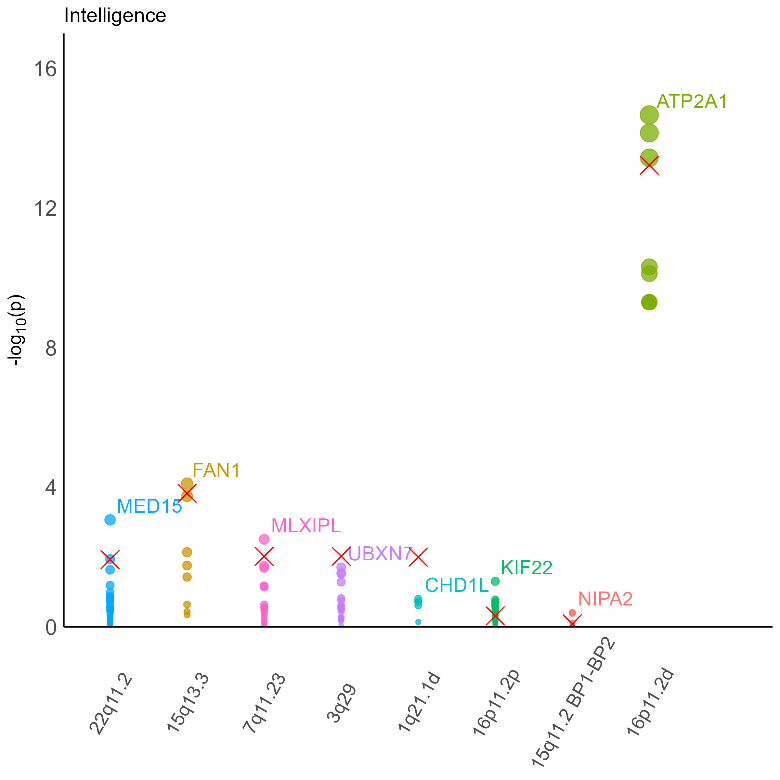 |
| 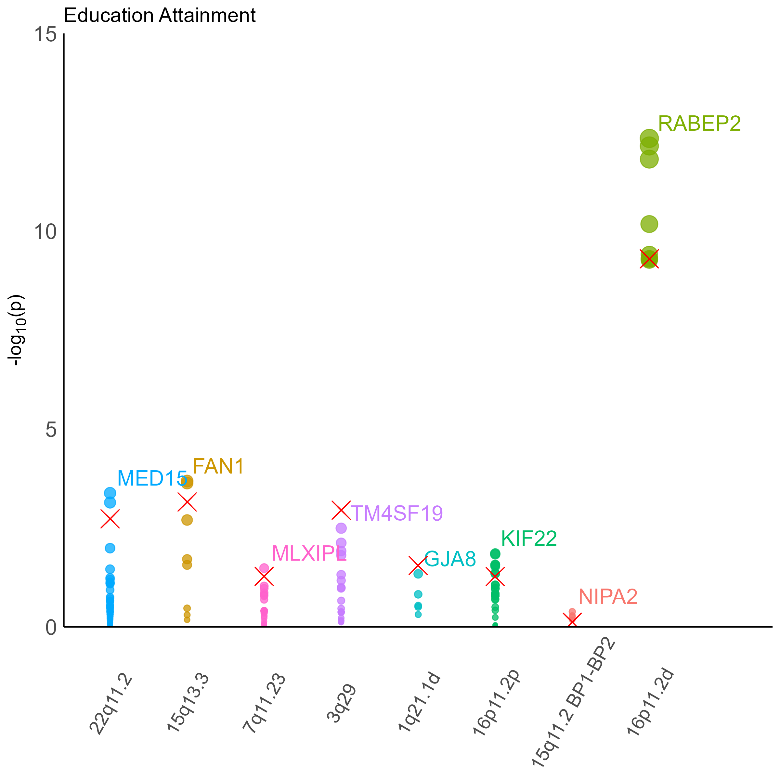 | 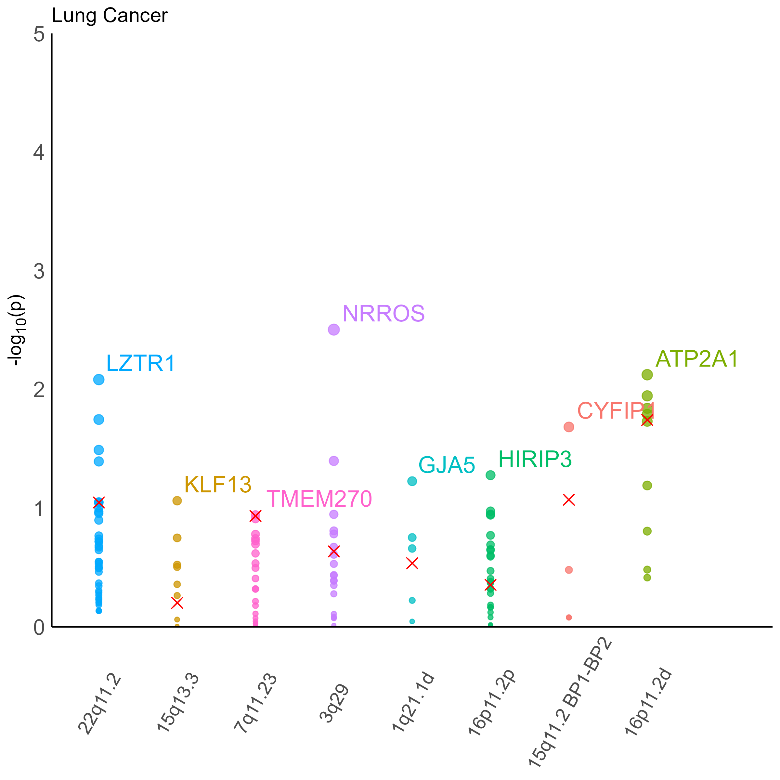 |
| 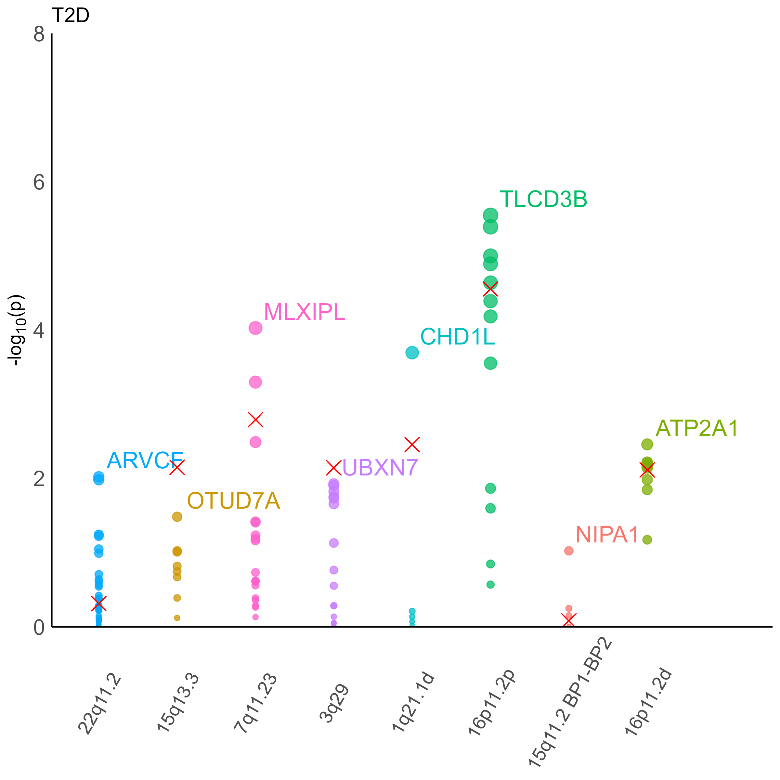 | 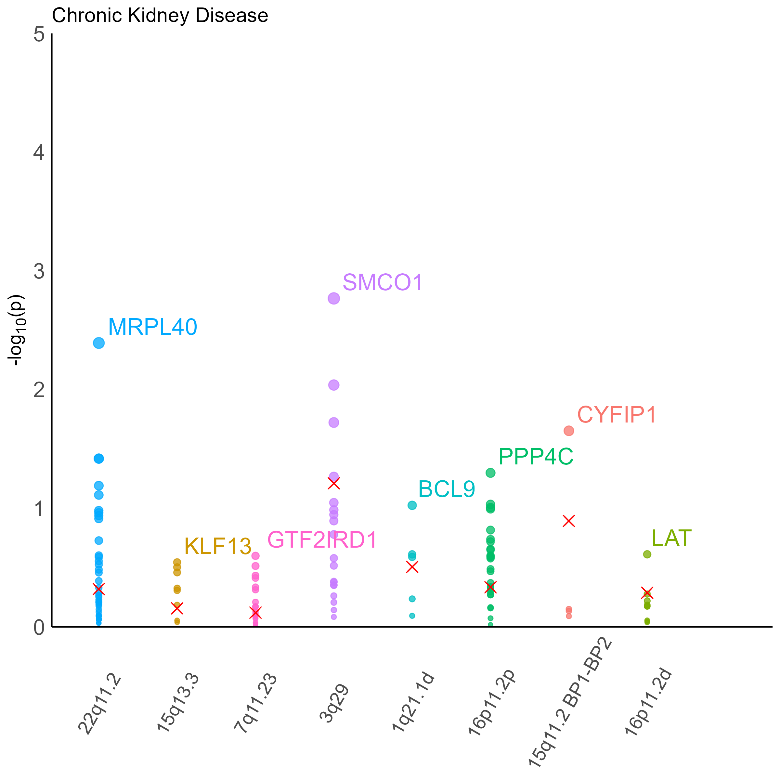 |
| 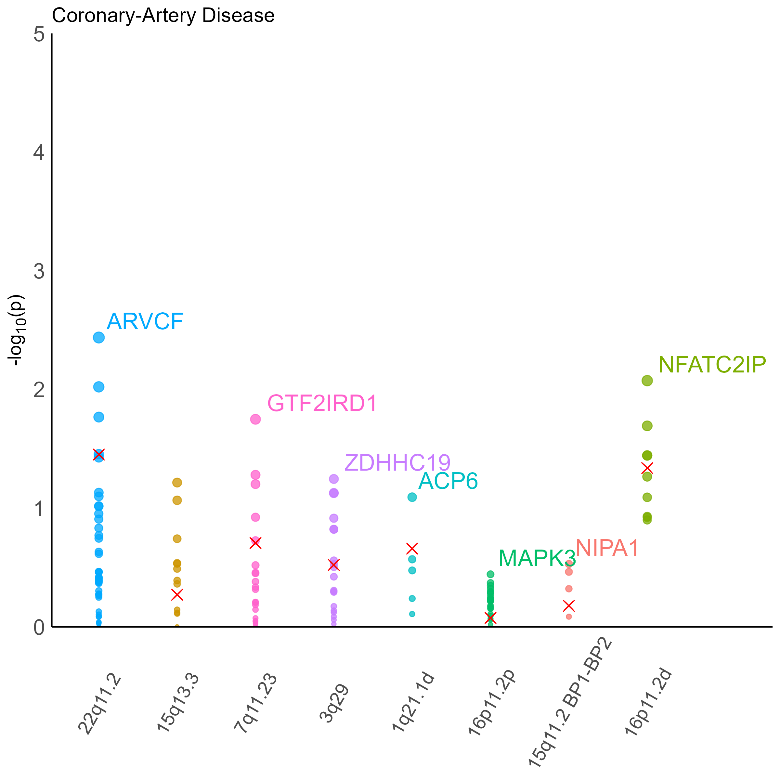 | 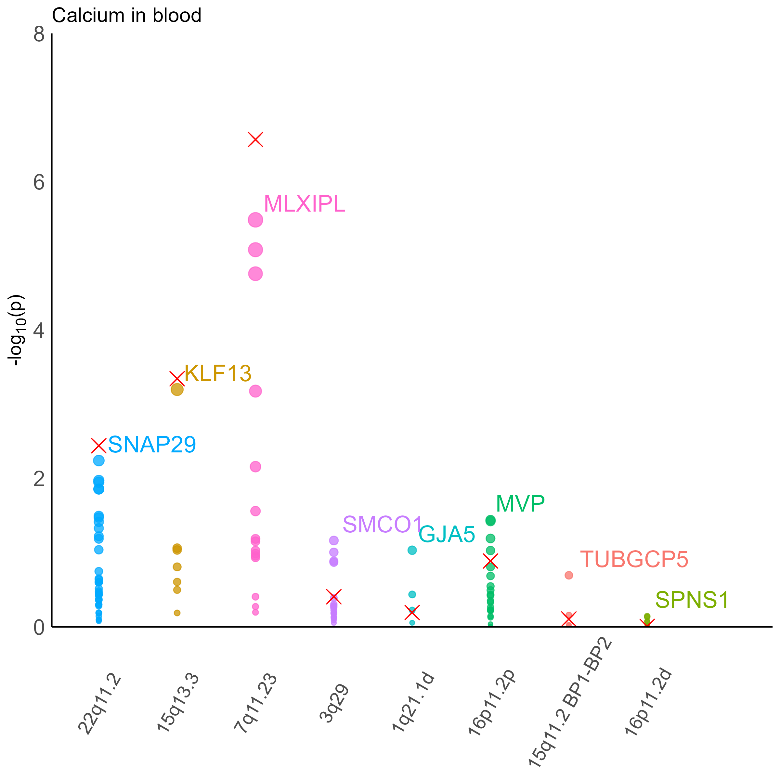 |
| 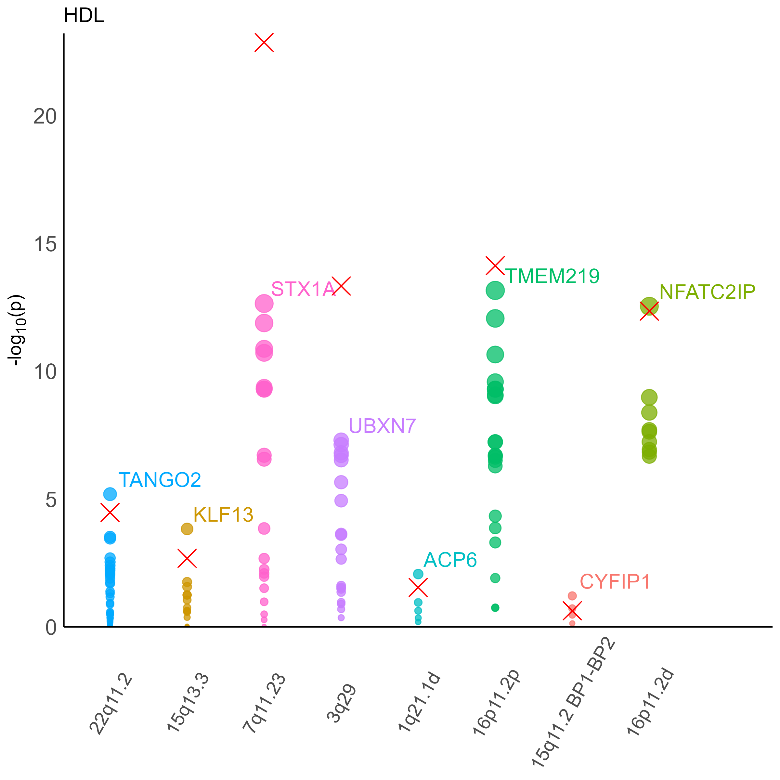 | 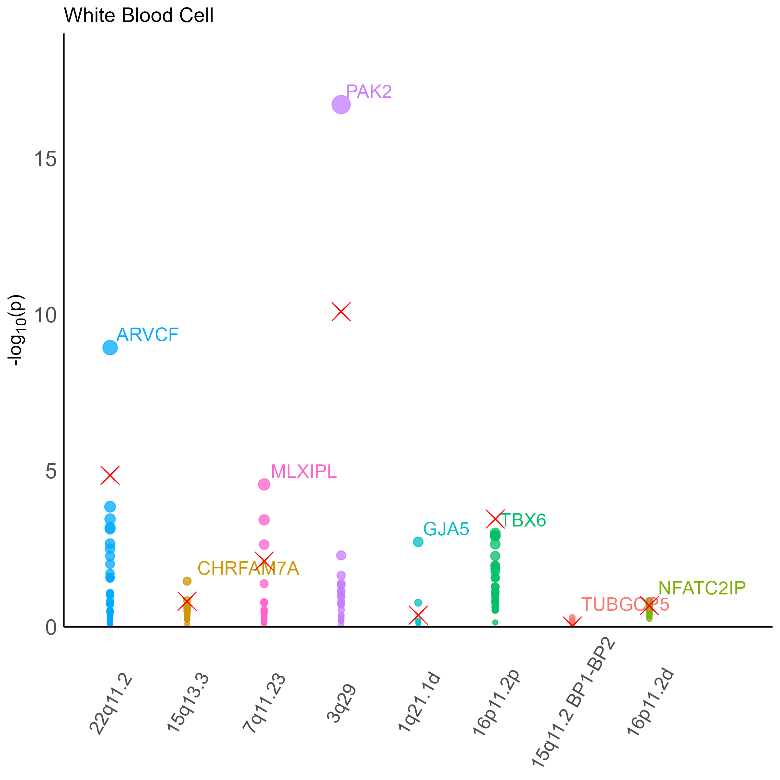 |
| 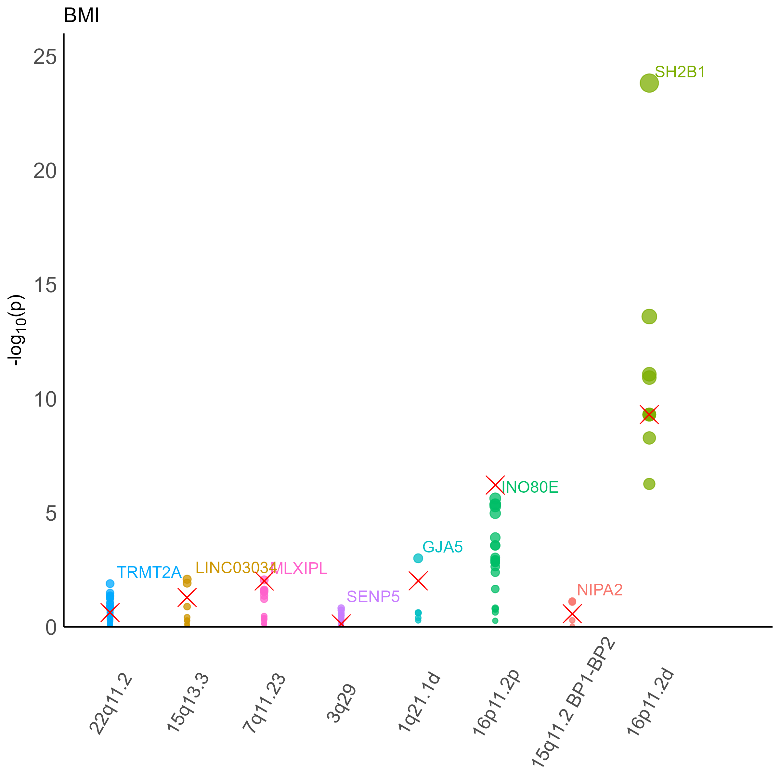 | 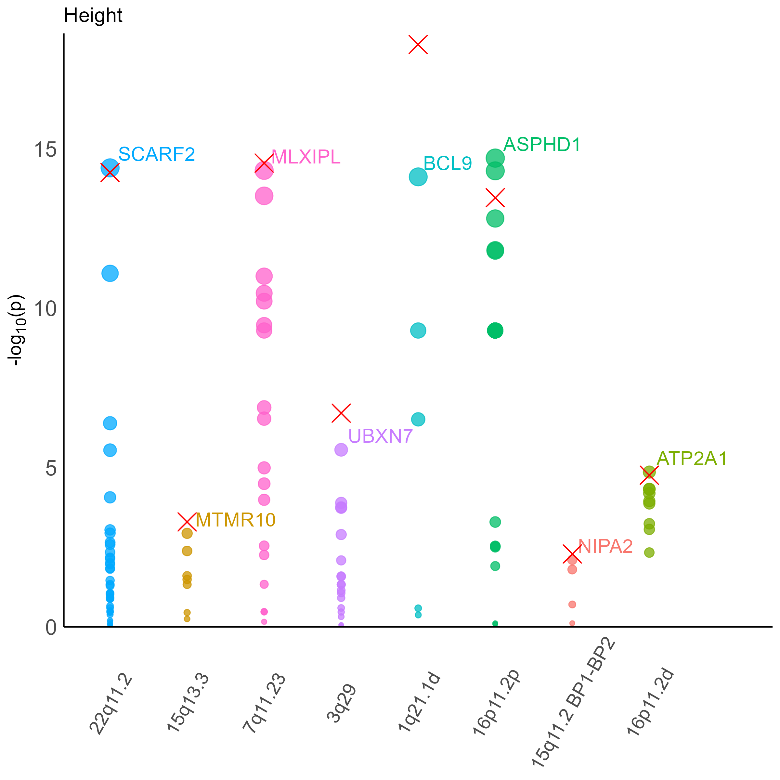 |

**Figure S5.** -log10 (p-values) for the individual genes in regions and the whole region of interest (cross) in MAGMA gene analysis for all 20 traits. The top gene is labelled. ADHD = Attention Deficit Hyperactivity Disorder, Alcohol = AUDIT score, ALZ = Alzheimer’s disease, ASD = Autism Spectrum Disorder, BMI = Body Mass Index, Cannabis = Cannabis Use Disorder, HDL = High-Density Lipoprotein, MDD = Major Depressive Disorder, PTSD = Post-Traumatic Stress Disorder, SCZ = schizophrenia, T2D = Type II Diabetes.
